# Social Determinants of Health and Chronic Disease Risk Prediction in the *All of Us* Research Program

**DOI:** 10.64898/2026.03.19.26348851

**Authors:** Matt Kammer-Kerwick, Yug Dave, Vidhi Parekh, Luke McDonald, S. Craig Watkins

## Abstract

Social determinants of health (SDoH), the social, economic, and environmental conditions shaping health trajectories, contribute to chronic disease risk comparably to clinical factors, yet most predictive studies model conditions independently, obscuring shared social pathways. Using participant-reported data from the *All of Us* Research Program (n=259,186), we evaluated the relative contributions of demographic factors and twelve SDoH domains to chronic disease prediction while accounting for the co-occurrence structure of conditions.

Hierarchical clustering identified two clinically meaningful outcome clusters: a Mental Health cluster (depression, anxiety, substance use disorder; prevalence = 51.7%) and a Cardiometabolic cluster (heart disease, diabetes, chronic lung disease; prevalence = 78.7%). Gradient boosted models were trained for each cluster under three feature configurations, SDoH only, demographics only, and combined, with performance evaluated using bootstrapped area under the receiver operating characteristic curve (AUC). Combined models achieved the highest discriminative performance for Mental Health (AUC = 0.701, 95% confidence interval: 0.696 - 0.705) and Cardiometabolic (AUC = 0.662, 95% CI: 0.655 - 0.668) outcomes. SDoH features outperformed demographics for Mental Health prediction (AUC = 0.678 vs. 0.655), while performance was comparable for Cardiometabolic outcomes (SDoH = 0.633; demographics = 0.636). Interpretability analysis using SHapley Additive exPlanations (SHAP) identified stress, discrimination, and religion/spirituality as the most influential SDoH domains for Mental Health outcomes; age, neighborhood disorder, and discrimination were primary predictors for Cardiometabolic outcomes. Double machine learning confirmed significant causal effects, with stress showing the largest average treatment effect on Mental Health outcomes (ATE = 0.093, p < 0.001). Interaction analyses revealed 24 significant SDoH-by-demographic interactions, indicating differential SDoH effects across racial/ethnic and gender/sexual minority subgroups.

These findings indicate that experiential social factors carry stronger predictive signal for mental health conditions, while Cardiometabolic conditions are more strongly shaped by demographic and structural neighborhood characteristics. Results support condition-specific SDoH screening protocols over universal instruments and targeted social interventions to reduce health disparities.

**Author Summary:** We developed and tested a four-stage analytical framework to predict chronic disease risk more precisely by combining individual Social Determinants of Health (one’s social environments, stress levels, neighborhood conditions, and community connections), with conventional patient demographics such as age, income, and race/ethnicity. Using data from nearly 260,000 participants in the *All of Us* Research Program, we found that including social and environmental factors meaningfully improve prediction of both mental health conditions (depression, anxiety, and substance use) and cardiometabolic conditions (heart disease, diabetes, and lung disease).

Importantly, not all social factors matter equally for all conditions. Mental health outcomes were most strongly shaped by experiential factors (stress, discrimination, and loneliness) while cardiometabolic outcomes were more strongly driven by age and neighborhood characteristics such as disorder and limited access to physical activity. We also found that stress, discrimination, and neighborhood disadvantage have stronger health effects among Black, Hispanic, and gender/sexual minority individuals, pointing to where targeted interventions could reduce persistent health disparities.

These findings suggest that clinicians and health systems should move away from one-size-fits-all social needs screening toward condition-specific tools that prioritize the social factors most relevant to the conditions being managed.

## 1 Introduction

The environments and social contexts in which individuals live from early life through older adulthood shape health trajectories long before clinical symptoms emerge [1]. These social determinants of health (SDoH) include upstream social and material conditions that influence exposure and vulnerability, contributing to persistent differences in health [2, 3]. Despite their importance, translating SDoH frameworks into routine risk assessment procedures remains challenging because clinical settings face pragmatic barriers related to workflow integration, documentation, and follow-up capacity [4, 5]. Moreover, evidence from prediction studies indicates that the marginal utility of predictive improvement from SDoH varies across outcomes and populations, highlighting the need for outcome-specific analytic strategies rather than a single SDoH assessment specification [6, 7].

Chronic conditions rarely occur in isolation. Multimorbidity, defined as the presence of multiple chronic conditions within an individual, has become a prevalent feature of population health [8]. Systematic reviews repeatedly identify stable multimorbidity profiles that distinguish mental-health clusters from Cardiometabolic clusters [9–11], providing a foundation for the clustering approach used in this study. When multiple outcomes are correlated, meaning they frequently co-occur, modeling each endpoint separately makes it difficult to interpret how predictors influence the underlying clinical processes they share. The clustering of correlated outcomes into clinically meaningful groups provides a more interpretable representation of these shared pathways and clarifies the contributions of each predictor [12].

In the present study, using data from the All of Us Research Program [13], we apply hierarchical clustering to eight chronic disease indications to characterize comorbidity structure and define clinically interpretable groupings for cross-indication comparison. To align predictive modeling with interpretable and policy-relevant inference, we employ a four-stage analytical design: (1) Gradient-boosted tree models with SHAP values to identify predictive SDoH factors; (2) Double Machine Learning for causal effect estimation; (3) Logistic regression for clinically interpretable odds ratios; and (4) Interaction analysis to identify differential SDoH effects across demographic subgroups. This multistage approach addresses limitations inherent in using any single analytical method and provides complementary perspectives on complex SDoH–health relationships [14, 15].

### 1.1 Theoretical Framework

The life course perspective, a well-established framework in social epidemiology, holds that upstream social and material conditions influence later disease risk through cumulative exposures that occur long before clinical presentation [1]. Structural determinants may function as fundamental causes by shaping access to flexible resources that can be deployed to avoid risks and minimize consequences of illness [16].

Multiple SDoH domains show robust associations with health outcomes across the empirical literature [3]. More advantaged socioeconomic position, including higher educational attainment and employment, is associated with decreased disease risk in multicohort studies [17], and higher income is strongly associated with longevity, with substantial geographic variation in the income–life expectancy gradient in the United States [18]. Stronger social relationships are associated with decreased mortality risk in meta-analytic evidence [19], and discrimination is associated with adverse mental and physical health outcomes [20]. In predictive modeling, scoping reviews of social and behavioral determinants in risk prediction report that the incremental utility of SDoH varies across outcomes, modeling contexts, and populations [6, 7]. Taken together, these findings highlight substantial heterogeneity in how different SDoH domains relate to different health outcomes, indicating that outcome-specific model development is needed rather than relying on a single SDoH specification to generalize across conditions.

Because many outcomes themselves co-occur rather than occurring independently, it is important to consider higher-level groupings when modeling social determinants. Multimorbidity prevalence increases with age in population cohorts [21], and systematic reviews frequently identify recurring groupings that include mental-health–related and cardiometabolic profiles [9–11]. These multimorbidity patterns provide the foundation for examining how demographic characteristics stratify exposure to social conditions and contribute to the clustering of health risks.

### 1.2 The Current Study

This research is guided by the following questions:

- **RQ1.** Which SDoH factors are most predictive of adverse health outcomes, and how do these predictors vary across different condition clusters?
- **RQ2.** What are the causal effects of SDoH factors on health outcomes after accounting for confounding, and do these effects exhibit heterogeneity across demographic subgroups?
- **RQ3.** Can SDoH factors be used to develop clinically interpretable risk stratification tools that provide actionable insights for intervention?
- **RQ4.** How do SDoH factors differentially impact health outcomes across demographic groups, and which

SDoH × demographic interactions are most clinically significant?

By examining these research questions, our objective is to extend beyond findings limited to specific conditions and develop a broader understanding of the ways in which social and demographic determinants jointly influence health outcomes. Although our selected domains are informed by existing scholarship, we recognize that additional social determinants may also have significant effects, and we openly address these limitations. Ultimately, our aim is to contribute to the development of more equitable and contextually informed approaches to disease prevention and risk assessment [13, 22, 23].

### 1.3 Analytical Framework

Relationships among SDoH domains and chronic health outcomes are high-dimensional, nonlinear, and characterized by correlated exposures and outcomes. No single analytic method can adequately support discovery, causal estimation, and clinically meaningful interpretation at the same time. We therefore implemented a four-stage analytical framework that integrates complementary approaches: machine learning for pattern discovery, causal inference for effect estimation, traditional biostatistics for clinically interpretable estimates, and formal interaction analysis for assessing differential impacts across demographic groups. This staged design mitigates the limitations inherent in any single method and provides a coherent, multi-perspective assessment of SDoH–health relationships. The complete analytical workflow is summarized in Fig 1.

**Fig 1:**
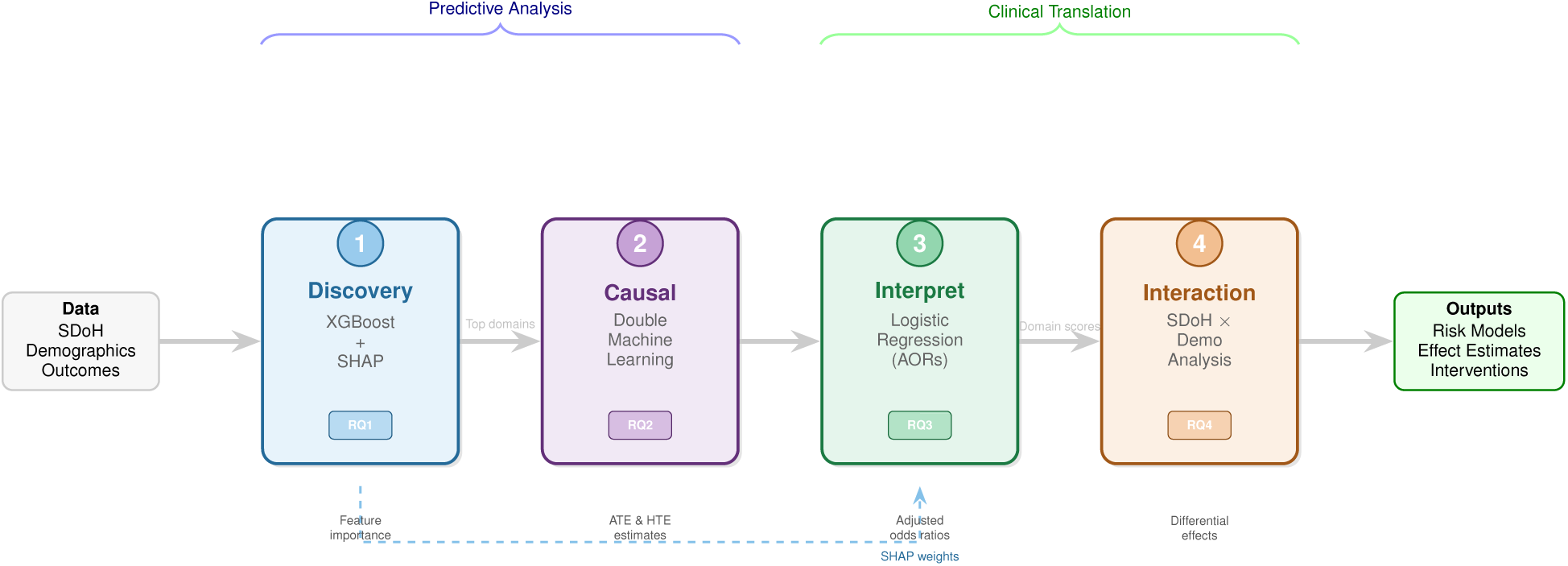
Four-Stage Analytical Framework for SDoH-Health Outcome Analysis. The pipeline progresses from predictive modeling (Stages 1–2) to clinical translation (Stages 3–4). Stage 1 uses XGBoost with SHAP to identify important SDoH predictors (RQ1). Stage 2 applies Double Machine Learning for causal effect estimation (RQ2). Stage 3 provides interpretable odds ratios via logistic regression (RQ3). Stage 4 tests differential effects across demographic groups (RQ4). The dashed arrow indicates that SHAP weights from Stage 1 inform domain score construction in Stage 3.

The four stages connect to our research questions as follows. Stage 1 (Discovery) uses gradient-boosted tree models with SHAP values to identify predictive SDoH factors and quantify domain-level importance (RQ1). Stage 2 (Causal Inference) uses Double/Debiased Machine Learning to estimate average and heterogeneous treatment effects across demographic subgroups (RQ2). Stage 3 (Interpretation) uses logistic regression with SHAP-weighted domain scores to estimate adjusted odds ratios with confidence intervals for clinical interpretation (RQ3). Stage 4 (Interaction Analysis) formally tests SDoH × demographic interactions to identify differential impacts (RQ4).

## 2 Materials and Methods

### 2.1 Data and Sample

Data were obtained from the All of Us Research Program, a national precision health initiative sponsored by the National Institutes of Health that aims to enroll one million or more participants reflecting the diversity of the United States [13]. The program collects electronic health records (EHR), survey responses, physical measurements, and biospecimens from participants across all 50 states. Our analysis used the Controlled Tier release, which provides access to linked survey and EHR data.

We restricted the analytical sample to participants who completed the Social Determinants of Health survey module, consented to EHR linkage, and had complete demographic information. Participants missing essential predictor variables or outcome indicators were excluded. The final sample comprised 259,186 participants with 88 SDoH features and complete data across all modeling stages.

### 2.2 Health Outcomes and Condition Clustering

Health outcomes were defined using self-reported condition items from survey responses, capturing conditions aligned with EHR-based chronic condition definitions. Eight conditions were extracted: depression (n = 133,684 cases), anxiety (n = 98,923), lung disease (n = 62,283), heart disease (n = 261,617),diabetes (n = 121,503), HIV (n = 2,753), schizophrenia (n = 11,582), and substance use disorder (n = 81,860).

Because chronic conditions frequently co-occur, modeling each outcome independently would obscure shared social pathways. We therefore applied hierarchical clustering to identify natural groupings based on co-occurrence patterns [24]. Pearson correlations were computed between all pairs of binary condition indicators and subjected to agglomerative hierarchical clustering using Ward’s minimum variance method [25], with distance computed as *d_ij_* = 1−*r_ij_*. Cluster assignments were determined by visual inspection of the dendrogram and clinical interpretability.

This procedure identified four clusters. One cluster grouped depression, anxiety, and substance use disorders, reflecting their well-documented comorbidity in clinical populations. A second cluster combined heart disease, diabetes, and lung disease, consistent with shared metabolic and inflammatory mechanisms. Schizophrenia and HIV emerged as separate singleton clusters, likely reflecting distinct etiological profiles and low prevalence. We retained the Mental Health cluster (prevalence = 51.7%) and Cardiometabolic cluster (prevalence = 78.7%) as primary outcomes for downstream modeling, as these contained sufficient cases and clinical coherence for multistage analysis.

### 2.3 Predictor Variables

Our predictor set comprised two tiers: demographic characteristics and SDoH domain scores. These tiers were modeled both separately and jointly to quantify the incremental predictive value of social context beyond conventional demographic information.

#### Demographics

Demographic variables were encoded as follows: age (continuous, in years; mean = 54.6, SD = 17.0); race/ethnicity (categorical: White [reference], Asian, Black, Hispanic, Other); gender and sexual identity (Cisgender Heterosexual Male [reference], Cisgender Heterosexual Female, Gender/Sexual Minority); household income (continuous); education level (ordinal 1–7 scale, mean = 5.74; also dichotomized as college degree vs. no college degree); and housing status (own/stable vs. rent/insecure). These characteristics provide essential context for equity-focused modeling because they reflect unequal exposure to social risks and unequal access to structural resources across the life course [26].

Age functions both as an indicator of biological aging and of accumulated social exposures [1]. Sex and gender influence health through intersecting biological and social pathways, with women showing higher prevalence of anxiety and depression and men experiencing elevated cardiovascular risk [27]. Race and ethnicity are social constructs reflecting lived experience and structurally patterned opportunity rather than biological difference [20, 28]. We include race/ethnicity to support equity-focused interpretation, including evaluating whether SDoH effects vary across groups and whether contextual factors interact with social identity in shaping risk. Income reflects stratified access to material resources and is strongly associated with longevity [18]. Education and employment are core indicators of socioeconomic position and are consistently associated with lower premature mortality risk [17].

#### Social Determinants of Health

SDoH variables were organized into twelve theoretically derived domains based on established frameworks in social epidemiology [22, 23] and drawn from the standardized *All of Us* SDoH survey instruments [29, 30]. The twelve domains and their item counts are: Social Cohesion (3 items), Neighborhood Disorder (13 items), Neighborhood Access to Physical Activity (7 items), Social Support (9 items), Loneliness (8 items), Discrimination (16 items), Food Insecurity (2 items), Transience (1 item), Home Problems (1 item), Stress (10 items), Religion/Spirituality (7 items), and English Proficiency (1 item).

Each domain captures a distinct facet of social context relevant to health. Social cohesion reflects community-level trust and collective efficacy [31]. Neighborhood disorder captures visible physical and social incivilities signaling threat and instability [32]. Neighborhood access reflects availability of safe spaces for physical activity [33]. Social support measures the availability of emotional, informational, or tangible assistance [19, 34]. Loneliness captures subjective isolation and its links to depression, anxiety, and inflammation [35]. Discrimination captures chronic psychosocial stress from unfair treatment based on social identity [20]. Food insecurity reflects uncertain access to food and its links to healthcare barriers [36]. Transience reflects housing instability and disrupted continuity of care [37]. Home problems capture substandard housing conditions contributing to harmful exposures [38]. Stress reflects perceived strain related to finances, caregiving, and safety, linked to allostatic load and multisystem dysregulation [39, 40]. Religion/spirituality captures coping strategies, meaning-making, and social connectedness [41]. English proficiency reflects language barriers associated with differences in care quality and outcomes [42, 43].

Domain scores were constructed using SHAP-weighted aggregation derived from Stage 1 results, so that items contributing more to prediction received greater weight in the composite score. This approach ensures that domain measures reflect both theoretical structure and empirical importance. For multi-item domains, internal consistency was assessed using Cronbach’s alpha. Most domains demonstrated acceptable reliability (*α* ≥ 0.70), with Social Support (*α* = 0.93), Discrimination (*α* = 0.93), Religion/Spirituality (*α* = 0.93), and Food Insecurity (*α* = 0.90) showing strong internal consistency. Neighborhood Disorder (*α* = 0.58) and Stress (*α* = 0.41) showed lower consistency, suggesting these constructs may be multidimensional.

### 2.4 Data Processing

We implemented a structured data preparation pipeline to harmonize demographic information, SDoH survey responses, and condition-specific outcomes into a unified participant-level dataset. The pipeline included cohort definition, survey restructuring, missing-value imputation, variable encoding, and train-test splitting. All preprocessing steps followed a strict zero-leakage protocol: imputation and scaling parameters were learned exclusively from the training subset and then applied to validation and test subsets. Class imbalance was handled by computing `scale_pos_weight` from training data only. The full workflow is summarized in Algorithm 1 (Box C in S1 Text).

### 2.5 Stage 1: Predictive Discovery (XGBoost + SHAP)

Stage 1 addresses RQ1 by identifying which SDoH factors are most predictive of each health outcome cluster. Traditional regression approaches assume linear relationships and may miss the complex interactions and non-linearities that characterize real-world SDoH–health relationships [44]. Gradient boosting can capture these complexities, while SHAP values provide interpretable feature importance measures grounded in cooperative game theory [45].

We trained separate XGBoost models [46] for each health outcome under three feature configurations: SDoH-only, demographics-only, and combined (SDoH + demographics). This design directly quantifies the incremental predictive value of social context beyond demographic information. Models were configured with `max_depth`=5, `learning_rate`=0.05, `subsample`=0.8, `colsample_bytree`=0.8, and `scale_pos_weight` adjusted for class imbalance. Early stopping with patience of 50 rounds was used to prevent overfitting. Model performance was evaluated using bootstrapped AUC distributions with 95% confidence intervals (100 resamples).

SHAP values were computed using the TreeSHAP algorithm for exact computation in polynomial time [47]. Global feature importance was quantified as the mean absolute SHAP value across all test samples. To facilitate clinical interpretation, individual item-level SHAP values were aggregated to domain-level importance scores:

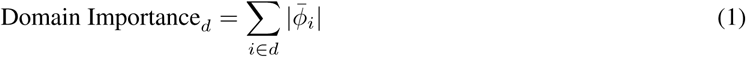

where *ϕ̄_i_* is the mean absolute SHAP value for item *i* belonging to domain *d*. These domain-level rankings informed the selection of focal exposures for subsequent stages. SHAP-derived weights for domain score construction were computed as:

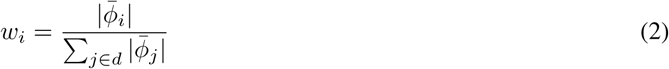

The modeling workflow is outlined in Algorithm 2 (Box C in S1 Text).

### 2.6 Stage 2: Causal Estimation (Double Machine Learning)

While Stage 1 identifies predictive associations, these may be confounded by demographic and socioeconomic factors. Stage 2 addresses RQ2 by estimating causal effects of SDoH factors on health outcomes using Double Machine Learning (DML) [48]. DML is well suited for settings where exposures are high-dimensional, correlated, and potentially confounded. Its key innovation is the use of Neyman orthogonality with cross-fitting, which reduces regularization and overfitting biases while allowing valid statistical inference for causal parameters.

For each outcome *Y* ∈ {0, 1} and focal SDoH domain *D* (selected from Stage 1 via SHAP importance), the Average Treatment Effect (ATE) was estimated as:

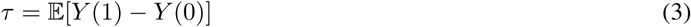

where *Y* (*d*) denotes the potential outcome under treatment level d. SDoH domain exposures entering DML were constructed using the same SHAP-weighted aggregation from Stage 1:

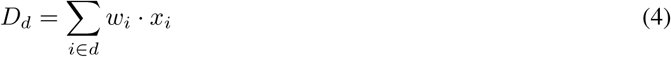

where *w_i_* are the SHAP-derived weights and *x_i_* are individual item responses. Domain scores were standardized to mean 0 and standard deviation 1. All twelve domains were evaluated as focal treatments, with remaining domains and demographic variables serving as confounders. The DML procedure is detailed in Algorithm 3 (Box C in S1 Text).

To assess whether SDoH effects varied across demographic subgroups, we also estimated conditional average treatment effects (CATEs) within strata defined by race/ethnicity and gender/sexual minority status. A minimum of 50 observations per subgroup was required for stable estimation.

### 2.7 Domain Selection for Clinical Modeling

Before proceeding to the clinical interpretation and interaction models in Stages 3 and 4, we conducted a structured domain review following Stages 1 and 2 to determine which SDoH domains should be carried forward. Not all twelve domains warrant inclusion in these later models. Retaining weakly predictive or causally null domains in logistic regression would reduce interpretability, increase the number of estimated parameters, and raise the risk of spurious findings. These concerns are particularly important in Stage 4, where each retained domain generates multiple interaction terms. Limiting the set of domains prior to fitting the clinical models therefore helps ensure that downstream interpretation focuses on the most substantively meaningful signals.

Because the retention decision required considering several complementary diagnostics, this filtering step was performed through researcher review rather than an automated selection procedure. The goal of the review was to evaluate the overall strength of evidence for each domain using information from the earlier modeling stages as well as measurement diagnostics. Each domain was evaluated using four criteria: (1) SHAP importance rank from Stage 1, indicating predictive contribution; (2) the double machine learning (DML) average treatment effect magnitude and statistical significance from Stage 2, indicating evidence of a causal association; (3) the variance inflation factor (VIF) derived from the domain correlation matrix, used to identify multicollinearity that could destabilize logistic regression estimates; and (4) Cronbach’s alpha, assessing whether the composite score exhibits sufficient internal consistency to support meaningful inference.

Based on these criteria, domains were classified into three tiers: *strong candidates* (high SHAP rank, statistically significant DML effect, and acceptable VIF), *review carefully* (mixed evidence across criteria), and *weak candidates* (low SHAP rank and non-significant or trivial DML effect). The complete domain review summaries, correlation matrices, and VIF tables are provided in Box B in S1 Text (Tables C and D). All domain retention decisions were made prior to fitting the Stage 3 and 4 models and are documented in the review tables provided in S1 Text.

For Mental Health, the five strong candidates were Stress (SHAP rank 1, ATE = 0.093), Discrimination (rank 2, ATE = 0.064), Religion/Spirituality (rank 3, ATE = −0.022), Loneliness (rank 4, ATE = 0.077), and Neighborhood Disorder (rank 5, ATE = 0.029). For Cardiometabolic outcomes, the five strong candidates were Neighborhood Disorder (rank 1, ATE = 0.026), Discrimination (rank 2, ATE = 0.023), Religion/Spirituality (rank 3, ATE = 0.005), Neighborhood Access (rank 4, ATE = 0.011), and Stress (rank 5, ATE = 0.036). All five strong candidates for each outcome showed significant DML effects (p *<* 0.001) and acceptable VIF values (≤ 1.6), providing converging evidence of both predictive and causal relevance.

The remaining seven domains were classified as *review carefully* or *weak candidates*. Social Support, for instance, ranked 6th–7th in SHAP importance but had a non-significant DML effect for Mental Health (ATE = 0.0005, p = 0.58), suggesting its predictive contribution may reflect confounding rather than a direct causal pathway. Social Cohesion similarly showed a null DML effect for Mental Health (ATE = 0.001, p = 0.27). Food Insecurity, while showing a significant DML effect for Mental Health, ranked 10th in SHAP importance and had a null effect for Cardiometabolic outcomes (ATE = 0.0004, p = 0.67). Transience was the sole weak candidate for Mental Health, ranking 11th with a non-significant DML effect (ATE = −0.001, p = 0.18). English Proficiency, Home Problems, and Transience are all single-item measures for which Cronbach’s alpha is not computable, further limiting confidence in their measurement precision.

Only the five strong candidate domains per outcome were carried forward into Stages 3 and 4. By restricting the clinical models to domains with converging evidence across complementary methods, high predictive importance, statistically significant causal effects, acceptable VIF levels, and adequate measurement reliability, we reduce the likelihood that noise-driven associations influence the interpretation stages and ensure that the interaction models in Stage 4 remain sufficiently focused to yield interpretable results.

### 2.8 Stage 3: Clinical Interpretation (Logistic Regression)

Stage 3 addresses RQ3 by translating the complex machine learning models into clinically interpretable risk estimates. While XGBoost achieves strong predictive accuracy, clinicians commonly prefer transparent models with easily communicated effect sizes [49]. Logistic regression provides Adjusted Odds Ratios (AORs) that are widely understood in clinical practice and medical research.

Multivariable logistic regression models were fit for each outcome cluster using only the five SDoH domains retained in the domain selection step described in the previous section. These retained SHAP-weighted domain scores were included alongside demographic covariates as predictors:

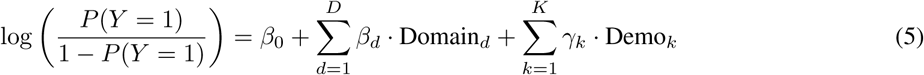

Domain scores were standardized (mean = 0, SD = 1), so AORs represent the multiplicative change in odds of the outcome per one standard deviation increase in the SDoH domain score, holding other variables constant. For Mental Health, the 5 strong candidate SDoH domains and 7 demographic variables were retained (12 predictors total); for Cardiometabolic outcomes, the 5 strong candidate SDoH domains and 6 demographic variables were retained (11 predictors total). Model discrimination was compared against the corresponding Stage 1 XGBoost models to quantify the interpretability–accuracy trade-off. The workflow is summarized in Algorithm 4 (Box C in S1 Text).

### 2.9 Stage 4: SDoH × Demographic Interaction Analysis

The preceding stages establish which SDoH domains predict and causally influence health outcomes on average across the full sample. But averages can obscure important variation in effect sizes across demographic groups. If stress exerts a stronger effect on cardiometabolic risk among Black and Hispanic participants than among White participants, a model that reports only the overall effect will understate risk for the former groups and overstate it for the latter. For health equity purposes, analysis of differential vulnerability assists clinicians and policymakers where screening and intervention will yield the greatest benefit and where universal approaches risk widening existing disparities [50].

Stage 4 addresses RQ4 by formally testing these SDoH × demographic interactions. Rather than testing all possible domain–modifier combinations, we restricted the interaction model to the five SDoH domains retained through the domain selection process described above. For Mental Health, these were Stress, Discrimination, Religion/Spirituality, Loneliness, and Neighborhood Disorder; for Cardiometabolic outcomes, Neighborhood Disorder, Discrimination, Religion/Spirituality, Neighborhood Access, and Stress. This design focuses on domains with the strongest prior evidence of predictive and causal relevance while keeping the total number of interaction terms manageable.

Demographic modifiers were chosen to represent the social identity dimensions most consistently linked to health disparities in the United States: race/ethnicity (indicator variables for Asian, Black, Hispanic, and Other, with White as reference) and gender/sexual minority status. These categories capture structurally patterned differences in exposure to discrimination, neighborhood disadvantage, and social support availability that are hypothesized to moderate SDoH– health relationships [28]. Extended logistic regression models included main effects for the 5 SDoH domains and 5 demographic modifiers, plus all 25 pairwise multiplicative interaction terms:

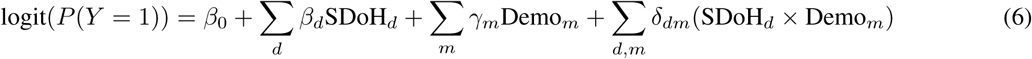

The interaction coefficient *δ_dm_* represents the difference in the log-odds of the outcome for domain *d* between the modifier and reference groups. A positive coefficient indicates that the SDoH effect is larger in the modifier group, while a negative coefficient indicates it is smaller. To make these effects clinically interpretable, we computed stratified AORs for each significant interaction. These stratified AORs represent the odds ratio for the SDoH domain estimated separately within the reference group (modifier = 0) and the modifier group (modifier = 1), allowing direct comparison effect magnitudes across populations.

Given the 50 interaction tests conducted across both outcomes (25 per outcome), we applied the Benjamini-Hochberg procedure to control the False Discovery Rate at 5% [51]. Both uncorrected and FDR-adjusted p-values are reported. Beyond statistical significance, we evaluated clinical significance by considering whether the interaction AOR exceeded conventional thresholds (> 1.5 or < 0.67), whether the stratified effects were consistent with known disparities mechanisms, and whether the interaction was biologically plausible. The workflow is summarized in Algorithm 5 (Box C in S1 Text).

### 2.10 Software and Reproducibility

All analyses were conducted using Python 3.10 in the All of Us Researcher Workbench. Key packages included: `pandas` (data manipulation), `numpy` (numerical computing), `scikit-learn` (machine learning utilities, including `train_test_split`, `StandardScaler`, `KFold`, `RidgeCV`), `xgboost` (gradient boosting), `shap` (SHAP value computation via `TreeExplainer`), `statsmodels` (logistic regression), `scipy` (hierarchical clustering, statistical tests), and `matplotlib`/`seaborn` (visualization). We interpret model performance and report validity considerations in line with guidance promoting complete and transparent reporting in clinical prediction modeling, including models that use machine learning methods [52]. Analysis code is available upon reasonable request to the corresponding author.

## 3 Results

### 3.1 Data Summary

The final analytical sample included 259,186 participants with complete data across all SDoH domains, demographic characteristics, and health outcome indicators. Demographic characteristics are presented in Table 1. The sample was predominantly White (69.6%), cisgender heterosexual female (57.4%), and college-educated (59.4%), with a mean age of 54.6 years (SD = 17.0).

**Table 1:**
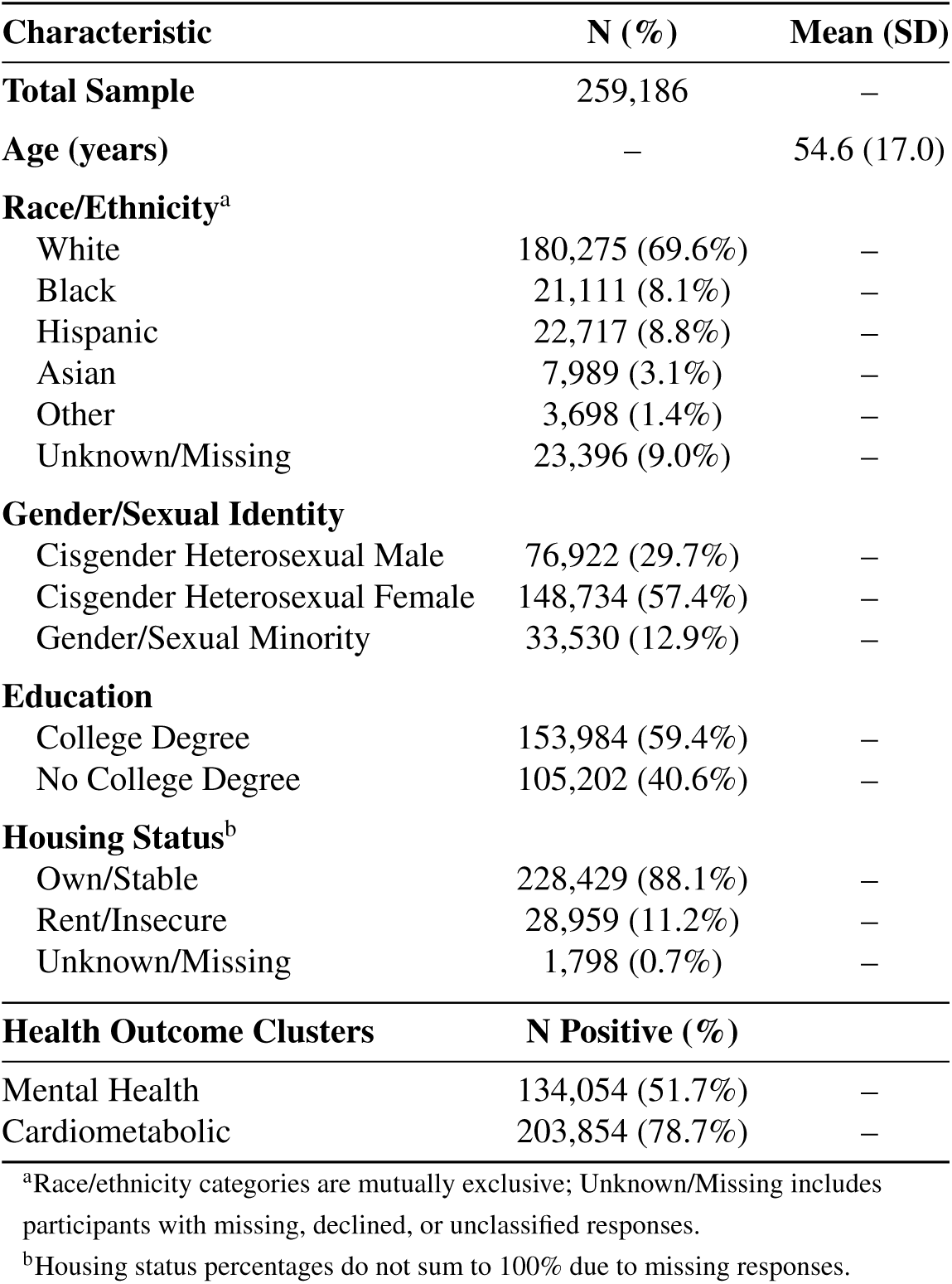
Demographic and Health Outcome Characteristics of Study Sample.

| Characteristic | N (%) | Mean (SD) |
| --- | --- | --- |
| **Total Sample** | 259,186 | – |
| **Age (years)** | – | 54.6 (17.0) |
| **Race/Ethnicity<sup>a</sup>** |  |  |
| White | 180,275 (69.6%) | – |
| Black | 21,111 (8.1%) | – |
| Hispanic | 22,717 (8.8%) | – |
| Asian | 7,989 (3.1%) | – |
| Other | 3,698 (1.4%) | – |
| Unknown/Missing | 23,396 (9.0%) | – |
| **Gender/Sexual Identity** |  |  |
| Cisgender Heterosexual Male | 76,922 (29.7%) | – |
| Cisgender Heterosexual Female | 148,734 (57.4%) | – |
| Gender/Sexual Minority | 33,530 (12.9%) | – |
| **Education** |  |  |
| College Degree | 153,984 (59.4%) | – |
| No College Degree | 105,202 (40.6%) | – |
| **Housing Status<sup>b</sup>** |  |  |
| Own/Stable | 228,429 (88.1%) | – |
| Rent/Insecure | 28,959 (11.2%) | – |
| Unknown/Missing | 1,798 (0.7%) | – |
| **Health Outcome Clusters** | **N Positive (%)** |  |
| Mental Health | 134,054 (51.7%) | – |
| Cardiometabolic | 203,854 (78.7%) | – |

<sup>a</sup>Race/ethnicity categories are mutually exclusive; Unknown/Missing includes participants with missing, declined, or unclassified responses.
<sup>b</sup>Housing status percentages do not sum to 100% due to missing responses.

We observed substantial co-occurrence across indications, consistent with population evidence that chronic conditions cluster due to shared determinants [8, 9]. Hierarchical clustering of the eight health conditions identified four distinct clusters (Fig A in S1 Text). The resulting groupings aligned with commonly observed multimorbidity profiles that distinguish mental health–related and Cardiometabolic patterns [10, 11, 53]. Schizophrenia (3.3% prevalence) and HIV (0.8%) emerged as separate singleton clusters. The prevalence of each retained outcome cluster is shown in the lower panel of Table 1.

### 3.2 Stage 1: Discovery Analysis (RQ1)

Table 2 summarizes predictive performance for XGBoost models across the three feature configurations. The Combined models (SDoH + Demographics) consistently achieved higher AUC values than single-domain models, demonstrating the incremental predictive value of SDoH beyond demographic information alone.

**Table 2:** Stage 1: Model Performance (Test Set AUC) Across Feature Configurations. Values represent median AUC with 95% bootstrap confidence intervals (100 resamples).

| Outcome | SDoH Only | Demo Only | Combined |
| --- | --- | --- | --- |
| Mental Health | 0.678 (0.673–0.681) | 0.655 (0.650–0.660) | 0.701 (0.696–0.705) |
| Cardiometabolic | 0.633 (0.626–0.638) | 0.636 (0.629–0.643) | 0.662 (0.655–0.668) |

Aggregating feature-level SHAP values to the domain level revealed distinct patterns of SDoH importance across outcome clusters (Fig 2, Table 3). For Mental Health outcomes, the Stress, Discrimination, and Religion/Spirituality domains showed the highest predictive importance. For Cardiometabolic outcomes, Age emerged as the strongest predictor, followed by Neighborhood Disorder and Discrimination. These domain-level importance rankings informed the selection of focal exposures for Stage 2 causal analysis.

**Fig 2:**
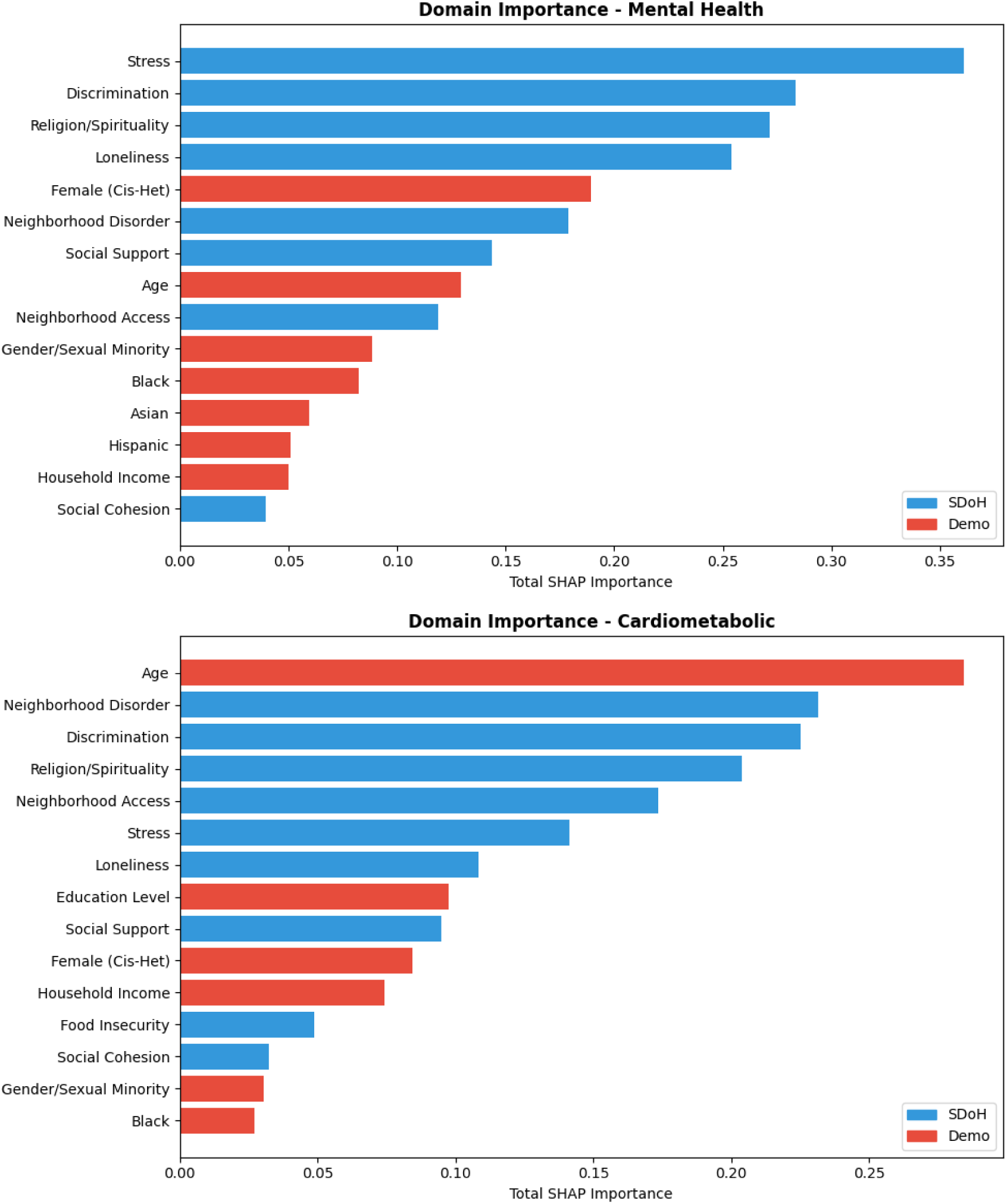
Domain-Level SHAP Importance Across Outcome Clusters. Aggregated SHAP importance by SDoH domain for the Mental Health (top) and Cardiometabolic (bottom) outcome clusters. Domain importance represents the sum of mean absolute SHAP values across all items within each domain.

**Table 3:** Stage 1: Domain-Level SHAP Importance by Outcome. Values represent total SHAP importance (sum of mean |SHAP| across items in domain). Top 3 domains per outcome are bolded.

| Domain | Mental Health | Cardiometabolic |
| --- | --- | --- |
| **Stress** | **0.361** | 0.141 |
| Loneliness | 0.254 | 0.108 |
| **Discrimination** | **0.283** | **0.225** |
| Social Support | 0.144 | 0.095 |
| **Neighborhood Disorder** | 0.179 | **0.232** |
| Food Insecurity | 0.017 | 0.049 |
| Social Cohesion | 0.039 | 0.032 |
| Neighborhood Access | 0.119 | 0.174 |
| Home Problems | 0.010 | 0.006 |
| **Religion/Spirituality** | **0.272** | 0.204 |
| Transience | 0.012 | 0.008 |
| English Proficiency | 0.032 | 0.004 |
| *Demo: Age* | 0.130 | **0.284** |
| *Demo: Income* | 0.050 | 0.074 |
| *Demo: Education* | 0.038 | 0.097 |

SHAP analysis of the Combined models identified the most predictive features for each outcome cluster (Fig B in S1 Text). Higher mean absolute SHAP values indicate greater feature importance, reflecting a larger average contribution of that feature to the model’s predicted probability of the outcome across individuals.

### 3.3 Stage 2: Causal Inference (RQ2)

Double Machine Learning was applied to estimate causal effects of all twelve SDoH domains on each outcome cluster. Table 4 presents the estimated Average Treatment Effects per one standard deviation increase in each SDoH domain, adjusted for demographic confounders. Stress exhibited the largest effect on both outcomes, with a one-standarddeviation increase associated with a 9.3 percentage-point increase in the probability of Mental Health outcomes (ATE = 0.093) and a 3.6 percentage-point increase in Cardiometabolic outcomes (ATE = 0.036). Loneliness (ATE = 0.077) and Discrimination (ATE = 0.064) also showed substantial effects on Mental Health.

**Table 4:** Stage 2: Average Treatment Effects (ATE) from Double Machine Learning. Values represent the estimated change in outcome probability per 1-SD increase in SDoH domain exposure, with 95% confidence intervals.

| SDoH Domain | Mental Health | Cardiometabolic |
| --- | --- | --- |
| Stress | 0.0931*** (0.0910–0.0949) | 0.0357*** (0.0339–0.0375) |
| Loneliness | 0.0769*** (0.0750–0.0788) | 0.0231*** (0.0212–0.0250) |
| Discrimination | 0.0635*** (0.0615–0.0655) | 0.0227*** (0.0207–0.0244) |
| English Speaking | 0.0356*** (0.0339–0.0376) | 0.0315*** (0.0296–0.0335) |
| Neighborhood Disorder | 0.0285*** (0.0269–0.0305) | 0.0255*** (0.0239–0.0271) |
| Social Support | 0.0005 (−0.0013–0.0024) | 0.0245*** (0.0233–0.0261) |
| Social Cohesion | 0.0011 (−0.0007–0.0029) | 0.0237*** (0.0219–0.0258) |
| Religion/Spirituality | −0.0219*** (−0.0239–−0.0202) | 0.0054*** (0.0040–0.0070) |
| Food Insecurity | 0.0201*** (0.0179–0.0223) | 0.0004 (−0.0017–0.0024) |
| Home Problems | 0.0165*** (0.0142–0.0185) | 0.0017 (−0.0002–0.0033) |
| Neighborhood Access | 0.0119*** (0.0099–0.0137) | 0.0112*** (0.0097–0.0128) |
| Transience | −0.0014 (−0.0034–0.0005) | −0.0073*** (−0.0089–−0.0056) |

\* p < 0.05; \*\* p < 0.01; \*\*\* p < 0.001

Several domains showed divergent patterns across outcomes. Religion/Spirituality was associated with a modest protective effect for Mental Health (ATE = −0.022) but a very small positive association with Cardiometabolic outcomes (ATE = 0.005). Social Support and Social Cohesion had non-significant effects on Mental Health but were significantly associated with Cardiometabolic outcomes. Food Insecurity was significant for Mental Health (ATE = 0.020) but not for Cardiometabolic outcomes.

Conditional ATEs estimated within demographic strata (Fig C in S1 Text) revealed notable heterogeneity. For example, the effect of Stress on Cardiometabolic outcomes was 1.20 times stronger among Hispanic participants compared to non-Hispanic participants. The effect of Loneliness on Mental Health showed significant variation by gender/sexual minority status, with effects being 0.96 times weaker in gender/sexual minority individuals.

### 3.4 Stage 3: Clinical Interpretation (RQ3)

Table 5 presents adjusted odds ratios from multivariable logistic regression models restricted to the five strong candidate SDoH domains per outcome, plus demographic covariates. The estimated effects are clinically interpretable and relevant for risk communication and clinical decision-making.

**Table 5:** Stage 3: Adjusted Odds Ratios from Logistic Regression. Models include only the five strong candidate SDoH domains per outcome, plus demographic covariates. AORs represent the multiplicative change in odds per 1-SD increase in standardized domain score.

| Domain | Mental Health<br>AOR [95% CI] | Cardiometabolic<br>AOR [95% CI] |
| --- | --- | --- |
| Stress | 1.37 (1.36–1.39)*** | 1.14 (1.13–1.15)*** |
| Discrimination | 1.11 (1.10–1.12)*** | 1.02 (1.01–1.03)*** |
| Religion/Spirituality | 0.93 (0.93–0.94)*** | 1.03 (1.02–1.04)*** |
| Loneliness | 1.19 (1.18–1.20)*** | – |
| Neighborhood Disorder | 1.03 (1.02–1.04)*** | 1.09 (1.08–1.10)*** |
| Neighborhood Access | – | 0.99 (0.98–1.00)* |
| Demo: Age | 1.06 (1.05–1.07)*** | 1.40 (1.38–1.41)*** |
| Demo: Female | 1.34 (1.33–1.35)*** | 1.13 (1.12–1.14)*** |
| Demo: Gender/Sexual Minority | 1.27 (1.25–1.28)*** | 1.06 (1.05–1.07)*** |
| Demo: Income | 1.09 (1.08–1.09)*** | 1.17 (1.16–1.18)*** |
| Demo: Education | – | 1.18 (1.17–1.19)*** |
| Demo: Black | 0.84 (0.83–0.84)*** | 0.92 (0.91–0.92)*** |
| Demo: Asian | 0.81 (0.80–0.82)*** | – |
| Demo: Hispanic | 0.84 (0.84–0.85)*** | – |
| **Model AUC** | 0.682 | 0.630 |
| **XGBoost AUC** | 0.701 | 0.662 |

\* p < 0.05; \*\* p < 0.01; \*\*\* p < 0.001

For Mental Health outcomes, Stress was the strongest SDoH predictor (AOR = 1.37), followed by Loneliness (AOR = 1.19) and Discrimination (AOR = 1.11). Religion/Spirituality was protective (AOR = 0.93), while Neighborhood Disorder showed a modest positive association (AOR = 1.03). Among demographic variables, cisgender heterosexual female sex (AOR = 1.34) and gender/sexual minority status (AOR = 1.27) were associated with elevated risk, while

Black (AOR = 0.84), Asian (AOR = 0.81), and Hispanic (AOR = 0.84) race/ethnicity were associated with lower risk relative to the White reference group. These patterns are illustrated in the forest plot in Fig 3, which highlights the dominant contribution of Stress and the protective association of Religion/Spirituality among SDoH domains, alongside the pronounced demographic effects of female sex and gender/sexual minority status.

**Fig 3:**
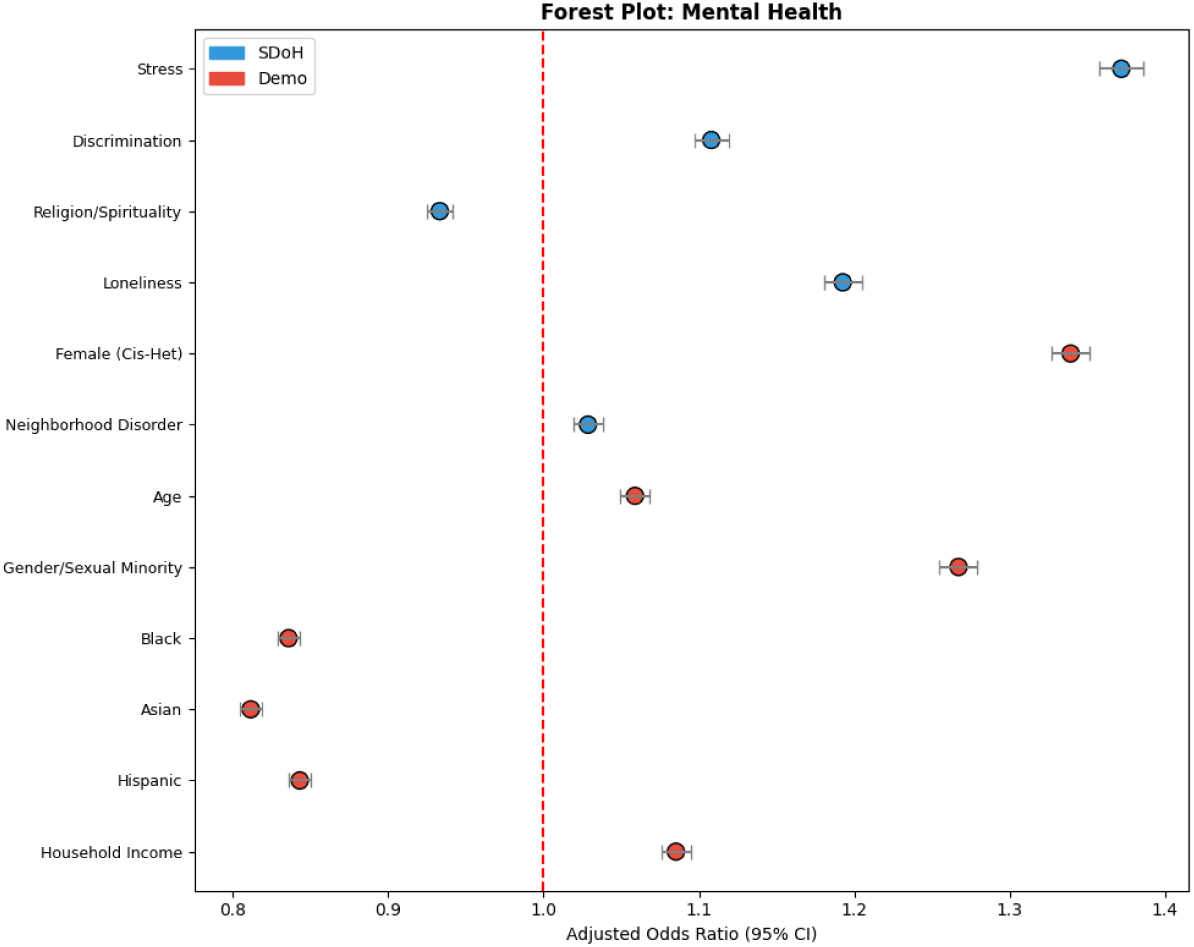
Forest plot of Adjusted Odds Ratios (AOR) with 95% confidence intervals for the Mental Health cluster. SDoH domains are shown in blue and demographic variables in red. The vertical dashed line at AOR=1 indicates no effect.

For Cardiometabolic outcomes, age was the dominant predictor (AOR = 1.40), reflecting the strong age-dependence of metabolic and cardiovascular disease. Among SDoH domains, Stress (AOR = 1.14) and Neighborhood Disorder (AOR = 1.09) showed the strongest associations. Religion/Spirituality (AOR = 1.03) and Discrimination (AOR = 1.02) had smaller but significant effects, while Neighborhood Access showed a borderline association (AOR = 0.99, p < 0.05). Education (AOR = 1.18) and income (AOR = 1.17) also contributed meaningfully. As shown in Fig 4, the outsized contribution of age relative to all SDoH domains distinguishes the Cardiometabolic risk profile from that of Mental Health, underscoring the greater role of biological aging and accumulated structural exposures in shaping cardiometabolic risk.

**Fig 4:**
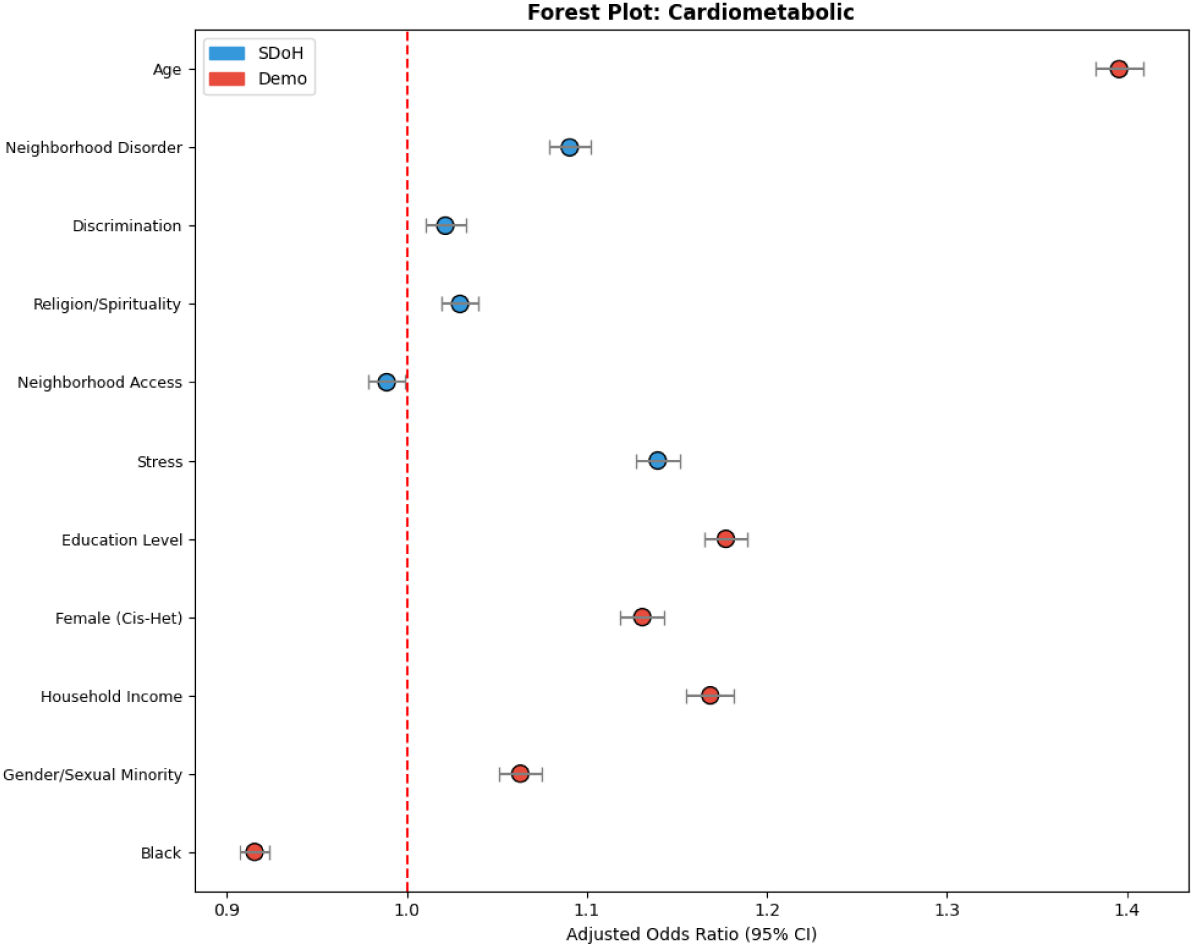
Forest plot of Adjusted Odds Ratios (AOR) with 95% confidence intervals for the Cardiometabolic cluster. SDoH domains are shown in blue and demographic variables in red. The vertical dashed line at AOR=1 indicates no effect.

Internal consistency of multi-item domain scores is reported in Table G in S1 Text. The logistic regression models achieved AUCs of 0.682 for Mental Health and 0.630 for Cardiometabolic outcomes, compared to 0.701 and 0.662 for the corresponding XGBoost models. The Mental Health model retained most of the XGBoost discriminative capacity (ΔAUC = 0.019), reflecting an acceptable trade-off for the gains in interpretability. The larger gap for Cardiometabolic outcomes (ΔAUC = 0.032) likely reflects the exclusion of domains such as Social Cohesion and Social Support that, while not meeting the converging-evidence threshold for strong candidacy, contributed predictive information in the more flexible XGBoost model.

### 3.5 Stage 4: Interaction Analysis (RQ4)

Extended logistic regression models were fit with main effects for the five retained SDoH domains and five demographic modifiers, plus all 25 pairwise interaction terms. Table 6 presents the main effects from the interaction models, which represent the estimated SDoH domain effects for the reference group (White, cisgender heterosexual participants). These reference-group effects differ from the Stage 3 estimates (Table 5) because the Stage 4 models include only the five demographic modifiers used in the interaction terms rather than the full set of demographic covariates, and because the inclusion of interaction terms redistributes shared variance between main effects and interaction coefficients. The interaction AORs in Table 7 are multiplicative modifiers of these reference-group baselines: the domain effect for a given subgroup is obtained by multiplying the main effect AOR by the corresponding interaction AOR. Model discrimination was 0.669 for Mental Health and 0.566 for Cardiometabolic outcomes.

**Table 6:** Stage 4: Main Effects (Reference-Group Baselines) from Interaction Models. AORs represent the SDoH domain effect for the reference group (White, cisgender heterosexual participants) per 1-SD increase in standardized domain score. These serve as the baseline against which the interaction AORs in Table 7 are interpreted.

| SDoH Domain | Mental Health<br>AOR [95% CI] | Cardiometabolic<br>AOR [95% CI] |
| --- | --- | --- |
| Stress | 1.41 (1.39–1.42)*** | 1.05 (1.03–1.06)*** |
| Discrimination | 1.10 (1.09–1.12)*** | 0.95 (0.94–0.97)*** |
| Religion/Spirituality | 0.96 (0.95–0.97)*** | 1.05 (1.03–1.06)*** |
| Loneliness | 1.18 (1.16–1.19)*** | – |
| Neighborhood Disorder | 1.02 (1.01–1.03)*** | 1.07 (1.05–1.08)*** |
| Neighborhood Access | – | 0.99 (0.98–1.00) |
| **Model AUC** | 0.669 | 0.566 |
| **Stage 3 AUC** | 0.682 | 0.630 |

\* p < 0.05; \*\* p < 0.01; \*\*\* p < 0.001
“–” indicates the domain was not retained for that outcome.

**Table 7:** Stage 4: Significant SDoH× Demographic Interactions. Interaction AORs represent the multiplicative modifier applied to the reference-group main effect (Table 6) for the indicated subgroup. The combined domain effect for a subgroup is: main effect AOR × interaction AOR. AOR *>* 1 indicates that the estimated domain–outcome association is shifted in the positive direction for the modifier group relative to the reference group. All *p*-values are FDR-adjusted.

| Outcome | Interaction Term | AOR | 95% CI | p-value |
| --- | --- | --- | --- | --- |
| Mental Health | Religion/Spirituality × Hispanic | 0.94 | 0.91–0.96 | <0.0001*** |
| Mental Health | Stress × Asian | 1.13 | 1.05–1.20 | 0.0005*** |
| Mental Health | Religion/Spirituality × Sexual Minority | 0.95 | 0.93–0.98 | 0.0007*** |
| Mental Health | Discrimination × Hispanic | 1.06 | 1.02–1.09 | 0.0019** |
| Mental Health | Loneliness × Sexual Minority | 0.96 | 0.93–0.98 | 0.0024** |
| Mental Health | Neighborhood Disorder × Black | 0.96 | 0.93–0.99 | 0.0058** |
| Mental Health | Religion/Spirituality × Asian | 0.93 | 0.88–0.98 | 0.0097** |
| Mental Health | Neighborhood Disorder × Hispanic | 0.97 | 0.94–1.00 | 0.0327* |
| Mental Health | Religion/Spirituality × Other | 0.93 | 0.87–1.00 | 0.0390* |
| Mental Health | Neighborhood Disorder × Other | 0.93 | 0.87–1.00 | 0.0417* |
| Cardiometabolic | Stress × Hispanic | 1.20 | 1.17–1.24 | <0.0001*** |
| Cardiometabolic | Stress × Black | 1.19 | 1.15–1.23 | <0.0001*** |
| Cardiometabolic | Neighborhood Access × Black | 1.11 | 1.07–1.15 | <0.0001*** |
| Cardiometabolic | Neighborhood Access × Hispanic | 1.10 | 1.06–1.14 | <0.0001*** |
| Cardiometabolic | Neighborhood Disorder × Black | 0.92 | 0.90–0.95 | <0.0001*** |
| Cardiometabolic | Religion/Spirituality × Black | 1.08 | 1.04–1.12 | <0.0001*** |
| Cardiometabolic | Stress × Asian | 1.12 | 1.06–1.19 | <0.0001*** |
| Cardiometabolic | Stress × Other | 1.18 | 1.09–1.28 | <0.0001*** |
| Cardiometabolic | Neighborhood Access × Sexual Minority | 1.07 | 1.03–1.10 | <0.0001*** |
| Cardiometabolic | Discrimination × Black | 1.06 | 1.03–1.10 | 0.0002*** |
| Cardiometabolic | Neighborhood Access × Asian | 1.10 | 1.03–1.16 | 0.0030** |
| Cardiometabolic | Neighborhood Disorder × Hispanic | 0.96 | 0.93–0.99 | 0.0060** |
| Cardiometabolic | Discrimination × Hispanic | 1.04 | 1.01–1.08 | 0.0192* |
| Cardiometabolic | Neighborhood Disorder × Other | 0.92 | 0.85–0.99 | 0.0327* |

\* p < 0.05; \*\* p < 0.01; \*\*\* p < 0.001 (FDR-adjusted)

Several notable shifts are apparent when comparing the reference-group main effects in the interaction model (Table 6) to the Stage 3 adjusted estimates (Table 5). For Cardiometabolic outcomes, the Stress main effect decreased from AOR = 1.14 in Stage 3 to AOR = 1.05 in the interaction model, while Discrimination shifted from AOR = 1.02 (positive) to AOR = 0.95 (negative). These shifts indicate that the Stage 3 pooled estimates partially reflected amplified effects in minoritized subgroups; once interactions are modeled explicitly, the reference-group baseline is lower. Similarly, for Mental Health, Stress increased from AOR = 1.37 to 1.41 in the interaction model, while Religion/Spirituality shifted from AOR = 0.93 to 0.96, reflecting the redistribution of variance once subgroup-specific effects are separated from the overall average.

Table 7 presents the significant interaction terms. For Mental Health, 10 significant interactions survived FDR correction; for Cardiometabolic outcomes, 14 did. The Mental Health interaction effects are visualized in Fig 5, here the most prominent patterns involve the amplified protective effect of Religion/Spirituality among Hispanic, Asian, and gender/sexual minority participants, and the notably stronger Stress effect among Asian participants. The corresponding Cardiometabolic interactions are shown in Fig 6, where the dominant feature is the amplified Stress effect across all minoritized racial/ethnic groups and the broadly stronger Neighborhood Access effects among Black, Hispanic, and Asian participants.

**Fig 5:**
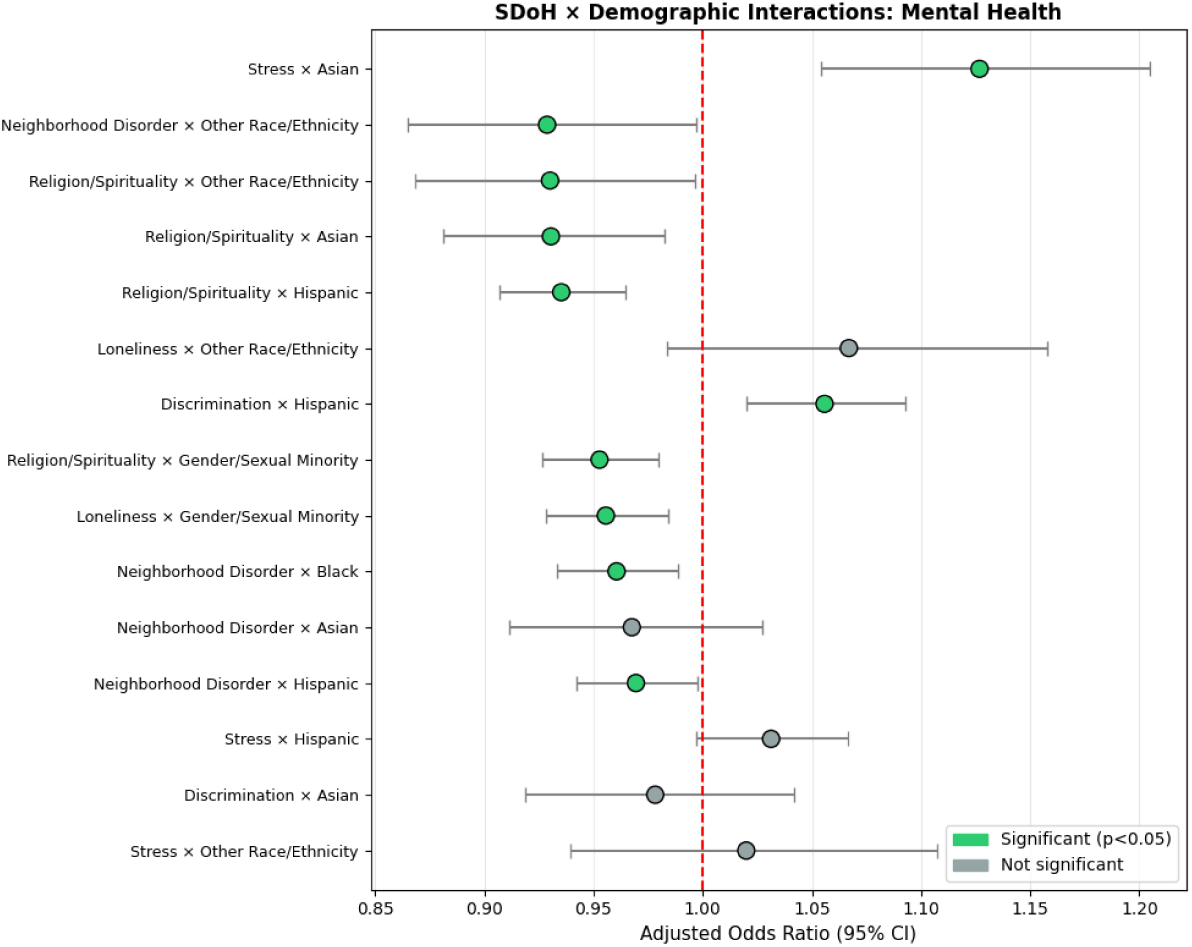
Forest plot of SDoH × Demographic interaction effects for the Mental Health cluster. Green points indicate statistically significant interactions (p *<* 0.05, FDR-adjusted).

**Fig 6:**
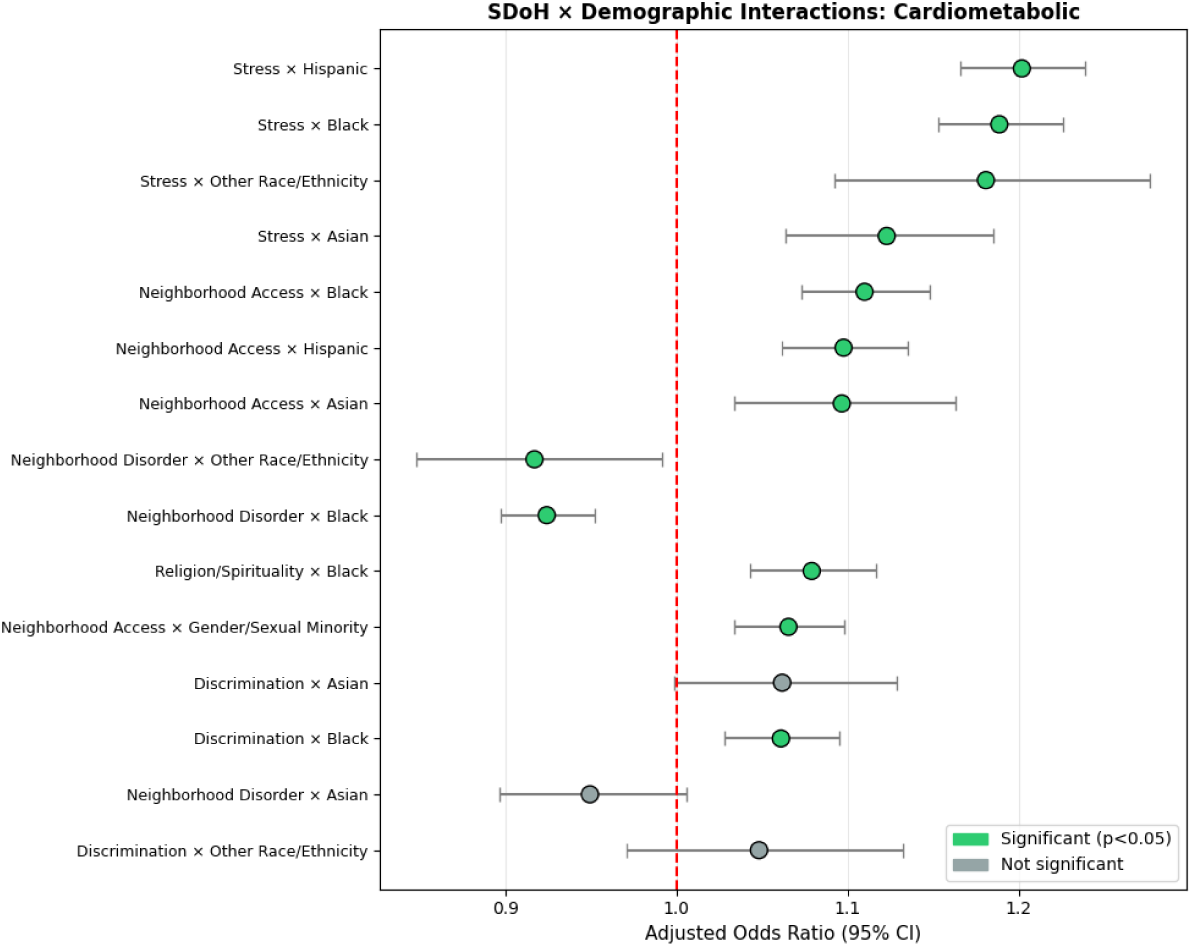
Forest plot of SDoH × Demographic interaction effects for the Cardiometabolic cluster. Green points indicate statistically significant interactions (p *<* 0.05, FDR-adjusted).

For Cardiometabolic outcomes, the most notable interactions involved Stress × race/ethnicity. The reference-group Stress effect was modest (AOR = 1.05; Table 6), but the Stress × Hispanic interaction AOR of 1.20 indicates a substantially amplified effect for Hispanic participants, yielding a combined subgroup effect of AOR = 1.05 × 1.20 = 1.26 (Table 8). Stress effects were similarly amplified among Black participants (interaction AOR = 1.19; combined AOR = 1.24), participants of Other race/ethnicity (interaction AOR = 1.18), and Asian participants (interaction AOR = 1.12). The contrast between the modest reference-group baseline and the amplified subgroup effects underscores that the Stage 3 pooled Stress estimate of AOR = 1.14 represented an average that understated risk for minoritized populations while overstating it for the reference group.

**Table 8:** Stage 4: Stratified Effects for Significant Interactions. AORs shown separately for the reference group (modifier = 0) and the modifier group (modifier = 1). The reference-group AOR corresponds to the main effect from Table 6; the modifier-group AOR is the product of the main effect and the interaction AOR from Table 7. The ratio indicates the fold-change in effect magnitude.

| Interaction | Outcome | AOR (Ref) | AOR (Mod) | Ratio |
| --- | --- | --- | --- | --- |
| Religion/Spirituality × Hispanic | Mental Health | 0.96 | 0.90 | 0.94× weaker |
| Stress × Asian | Mental Health | 1.41 | 1.59 | 1.13× stronger |
| Religion/Spirituality × Sexual Minority | Mental Health | 0.96 | 0.92 | 0.95× weaker |
| Discrimination × Hispanic | Mental Health | 1.10 | 1.16 | 1.06× stronger |
| Loneliness × Sexual Minority | Mental Health | 1.18 | 1.13 | 0.96× weaker |
| Stress × Hispanic | Cardiometabolic | 1.05 | 1.26 | 1.20× stronger |
| Stress × Black | Cardiometabolic | 1.05 | 1.24 | 1.19× stronger |
| Neighborhood Access × Black | Cardiometabolic | 0.99 | 1.10 | 1.11× stronger |
| Neighborhood Access × Hispanic | Cardiometabolic | 0.99 | 1.09 | 1.10× stronger |
| Neighborhood Disorder × Black | Cardiometabolic | 1.07 | 0.99 | 0.92× weaker |

Neighborhood Access showed differential effects for Cardiometabolic outcomes across multiple groups. The reference-group main effect was null (AOR = 0.99), but effects were 1.11-fold stronger among Black participants, 1.10-fold stronger among Hispanic participants, 1.10-fold stronger among Asian participants, and 1.07-fold stronger among gender/sexual minority individuals, potentially reflecting structural disparities in neighborhood investment where limited access to healthcare, healthy food, and recreational facilities disproportionately impacts marginalized communities. Conversely, Neighborhood Disorder showed weaker effects relative to the reference-group baseline (AOR = 1.07) among Black participants (0.92-fold; combined AOR = 0.99), Hispanic (0.96-fold), and Other (0.92- fold) participants, suggesting possible adaptation or resilience factors not captured in the model. Religion/Spirituality effects were amplified among Black participants for Cardiometabolic outcomes (1.08-fold), and Discrimination effects were stronger among Black (1.06-fold) and Hispanic (1.04-fold) participants.

For Mental Health, the reference-group Religion/Spirituality effect was modestly protective (AOR = 0.96; Table 6), but this protective association was amplified among Hispanic (interaction AOR = 0.94; combined AOR = 0.90), gender/sexual minority (interaction AOR = 0.95; combined AOR = 0.92), Asian (interaction AOR = 0.93), and Other race/ethnicity (interaction AOR = 0.93) participants. Stress effects on Mental Health, already strong in the reference group (AOR = 1.41), were further amplified among Asian participants (interaction AOR = 1.13; combined AOR = 1.59). Discrimination effects were more positive among Hispanic participants (interaction AOR = 1.06; combined AOR = 1.16 vs. reference AOR = 1.10). Loneliness effects were attenuated among gender/sexual minority individuals (interaction AOR = 0.96; combined AOR = 1.13 vs. reference AOR = 1.18), and Neighborhood Disorder effects were weaker among Black (0.96-fold), Hispanic (0.97-fold), and Other (0.93-fold) participants. Stratified effects for all significant interactions are presented in Table 8.

To illustrate the practical magnitude of interaction effects, Fig 7 presents predicted probability plots for the most clinically significant interactions. These plots display the model-estimated probability of the outcome as a function of the SDoH domain score, stratified by demographic group. Diverging slopes between the reference and modifier groups indicate differential vulnerability: steeper slopes in the modifier group reflect amplified SDoH effects, while converging slopes indicate attenuation. The predicted probability framework complements the AOR-based interaction estimates by placing effect modification on the absolute risk scale, which is more directly relevant for clinical decision-making and resource allocation.

**Fig 7:**
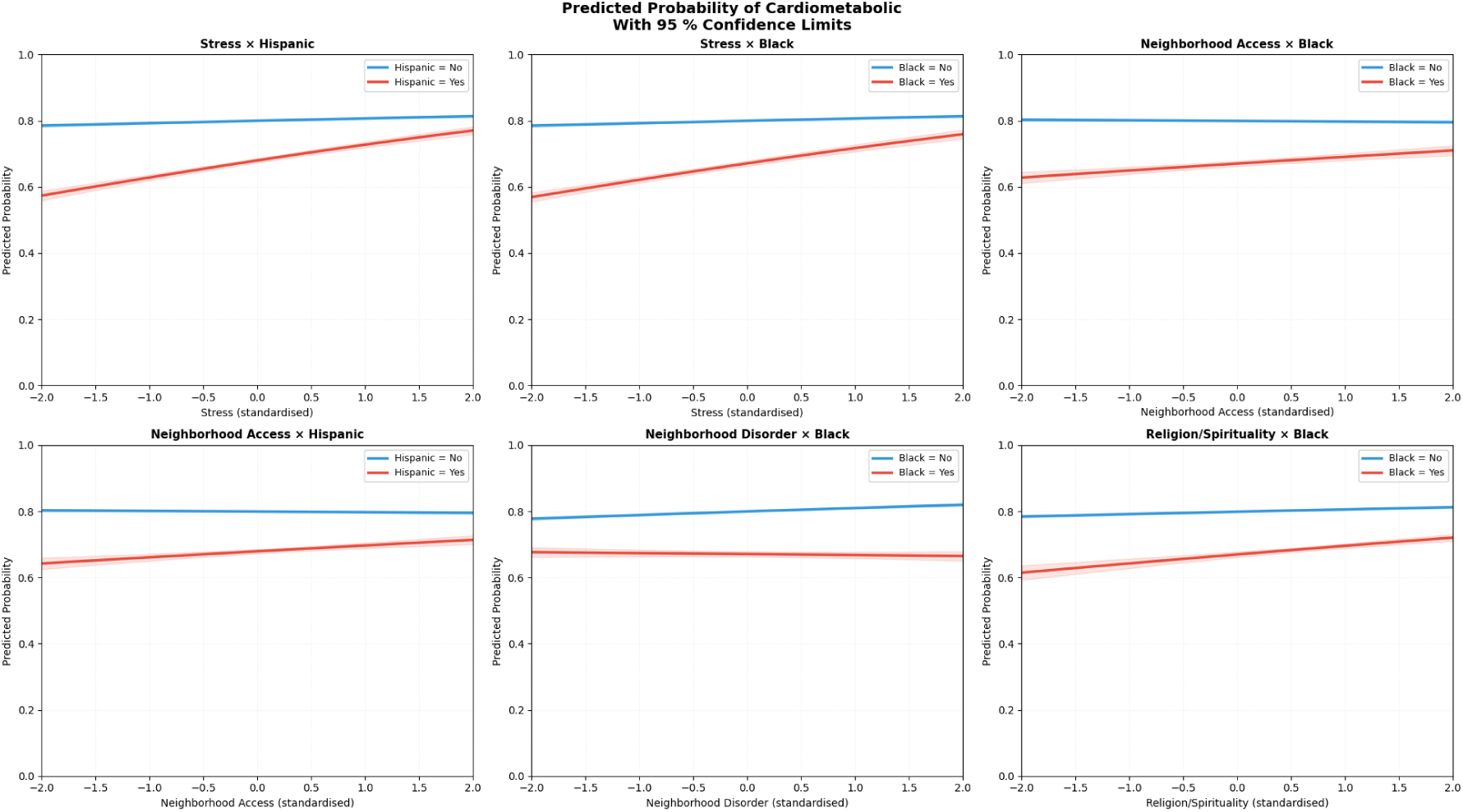
Predicted Probability Interaction Plots for Key SDoH × Demographic Interactions. Model-estimated probability of the outcome as a function of SDoH domain score, stratified by demographic modifier group. Diverging slopes indicate differential SDoH effects across subgroups. For Cardiometabolic outcomes, the steeper slopes for Hispanic and Black participants on the Stress domain illustrate the amplified stress–cardiometabolic association in these populations (interaction AOR = 1.20 and 1.19, respectively). For Mental Health outcomes, the steeper slope for Asian participants on Stress (interaction AOR = 1.13) and the enhanced protective effect of Religion/Spirituality among Hispanic participants (interaction AOR = 0.94) are shown. Shaded regions represent 95% confidence intervals.

### 3.6 Summary of Key Findings

Table 9 integrates findings across all four stages. Key findings include:

1. **Stage 1 (RQ1):** SDoH factors, particularly Stress (SHAP = 0.361), Discrimination (SHAP = 0.283), and Religion/Spirituality (SHAP = 0.272), were consistently among the strongest predictors of Mental Health outcomes. Combined models outperformed single-domain models across both outcome clusters, with AUC improvements of 0.023–0.046 over SDoH-only models and 0.026–0.046 over demographics-only models.
2. **Stage 2 (RQ2):** DML confirmed significant causal effects of multiple SDoH domains after adjusting for demographic confounders. Stress showed the largest causal effect on Mental Health (ATE = 0.093, 95% CI: 0.091–0.095) and Cardiometabolic (ATE = 0.036, 95% CI: 0.034–0.038) outcomes. Religion/Spirituality demonstrated a protective effect for Mental Health (ATE = −0.022, 95% CI: −0.024–−0.020).
3. **Stage 3 (RQ3):** Logistic regression models restricted to strong candidate domains demonstrated adequate discriminative performance while providing interpretable adjusted odds ratios. The Mental Health model retained most of the XGBoost capacity (ΔAUC = 0.019), while the Cardiometabolic model showed a larger gap (ΔAUC = 0.032) reflecting the exclusion of domains that contributed predictive but not causal signal. The strongest associations were observed for Stress (AOR = 1.37) and Loneliness (AOR = 1.19) with Mental Health, and Age (AOR = 1.40) with Cardiometabolic outcomes.
4. **Stage 4 (RQ4):** Formal interaction testing identified 10 significant SDoH × demographic interactions for Mental Health and 14 for Cardiometabolic outcomes after FDR correction. The strongest interactions involved Stress × Race/Ethnicity for Cardiometabolic outcomes, with effects 1.19–1.20 times stronger among Black and Hispanic participants. Predicted probability plots (Fig 7) confirmed that these interaction effects translate into clinically meaningful differences in absolute risk across demographic subgroups.

**Table 9:** Summary of Key Findings Across Four Stages.

| Outcome | XGBoost AUC | LR AUC | Top Causal Domain | N Sig. Interactions |
| --- | --- | --- | --- | --- |
| Mental Health | 0.701 | 0.682 | Stress | 10 |
| Cardiometabolic | 0.662 | 0.630 | Stress | 14 |

## 4 Discussion

### 4.1 Summary of Findings

We examined how demographics and 12 SDoH domains contribute to predicting multiple chronic disease indications in the All of Us cohort (n = 259,186). Using outcome clustering prior to cross-indication modeling, we observed groupings consistent with established multimorbidity patterns, including a Mental Health cluster (depression, anxiety, substance use disorder; prevalence = 51.7%) and a Cardiometabolic cluster (heart disease, diabetes, chronic lung disease; prevalence = 78.7%). These clusters align with groupings identified in systematic reviews of multimorbidity profiles [9–11] and replicate at a larger scale the mental health–cardiometabolic distinction documented in longitudinal urban cohorts [12]. Across indications, the combined SDoH+demographics models achieved the best predictive performance (Mental Health AUC = 0.701; Cardiometabolic AUC = 0.662), indicating that social context provides information that is complementary to demographic structure rather than redundant. These results extend our prior single-indication analysis of major depressive disorder in the All of Us dataset [53] to a multi-indication framework, confirming that the contribution of social determinants to health prediction generalizes beyond any individual condition.

### 4.2 Domain- and Outcome-Specific SDoH Contributions

A central finding of this study is that the predictive importance of SDoH varies substantially across domains and health outcomes. Stress (SHAP = 0.361), Discrimination (SHAP = 0.283), and Religion/Spirituality (SHAP = 0.272) emerged among the strongest SDoH predictors for Mental Health outcomes in Stage 1, with Loneliness (SHAP = 0.254) also demonstrating substantial predictive importance. In contrast, Cardiometabolic outcomes showed greater dependence on demographic characteristics, with age as the dominant predictor (SHAP = 0.284), followed by Neighborhood Disorder (SHAP = 0.232) and Discrimination (SHAP = 0.225). This pattern is also reflected in model performance: SDoH-only models outperformed demographics-only models for Mental Health (AUC 0.678 vs. 0.655), whereas the reverse was observed for Cardiometabolic outcomes (AUC 0.633 vs. 0.636).

The prominence of experiential social factors for mental health is consistent with a substantial body of evidence linking perceived stress to depression and anxiety through neuroendocrine dysregulation and allostatic load [39], and linking discrimination to adverse mental health outcomes through chronic psychosocial stress [20]. The strong predictive contribution of loneliness aligns with meta-analytic evidence that subjective social isolation is robustly associated with depressive symptomatology [19, 35]. Notably, our prior analysis of the All of Us dataset focusing on major depressive disorder similarly identified food insecurity and loneliness as strong predictors [53], and the current multi-indication framework confirms that these experiential domains extend their influence to the broader Mental Health cluster.

In contrast, the greater dependence of Cardiometabolic outcomes on age and structural neighborhood characteristics is consistent with evidence that cardiovascular and metabolic diseases reflect cumulative biological aging alongside long-term environmental exposures [1, 21]. The strong contribution of Neighborhood Disorder to Cardiometabolic prediction aligns with research linking visible neighborhood incivilities to physiological stress responses and reduced physical activity [32, 33].

These findings suggest that condition-specific SDoH assessment may be more actionable than applying a single screening approach across all conditions. Mental health outcomes appear particularly sensitive to experiential social factors that may be amenable to psychosocial interventions, whereas cardiometabolic outcomes may benefit more from upstream policy interventions targeting the built environment and economic security. This distinction has practical implications for the design of SDoH screening tools in clinical settings, where workflows already face pragmatic barriers [4, 5].

### 4.3 Causal Inference Findings

The Stage 2 DML analysis provided evidence that several SDoH domains exert causal effects on health outcomes beyond predictive association. Experiential social determinants, including stress, loneliness, and discrimination, emerged as the strongest causal contributors to Mental Health outcomes. Stress showed the largest estimated effect (ATE = 0.093), followed by loneliness (ATE = 0.077) and discrimination (ATE = 0.064). These magnitudes are substantively meaningful: a one-standard-deviation increase in the stress domain is associated with a 9.3 percentage-point increase in the probability of Mental Health cluster membership. These findings resonate with the allostatic load framework, which posits that chronic stress leads to multisystem physiological dysregulation with downstream mental health consequences [39, 40].

Religion/Spirituality demonstrated a modest protective effect for Mental Health (ATE = −0.022), consistent with systematic reviews linking religious engagement to improved psychological well-being through social support provision, coping resources, and existential meaning-making [41, 54]. Interestingly, this protective association was specific to Mental Health; Religion/Spirituality showed a small positive association with Cardiometabolic outcomes (ATE = 0.005), potentially reflecting confounding with age or sedentary social gathering patterns that were not fully captured by the DML adjustment.

For Cardiometabolic outcomes, the estimated causal effects for SDoH were generally smaller. Stress (ATE = 0.036), Neighborhood Disorder (ATE = 0.026), and Social Support (ATE = 0.025) emerged as the most influential domains, but with magnitudes substantially below those observed for Mental Health. This pattern suggests that while social determinants do causally influence cardiometabolic health, their effects may operate through longer-term behavioral and biological pathways, such as chronic inflammation, health behavior patterns, and healthcare access, that are only partially captured in cross-sectional analyses.

Several domains with the largest estimated causal effects also ranked among the most important predictors in the Stage 1 machine learning models. While SHAP-based weights were used to construct domain scores for DML, the causal estimation is orthogonalized and cross-fitted, so high predictive importance does not mechanically guarantee a large ATE. This empirical alignment between predictive discovery and causal estimation suggests that the signals captured by XGBoost correspond closely to domains with meaningful causal influence, providing greater confidence in the substantive importance of these domains than either method alone would offer.

### 4.4 Clinical Interpretability

Stage 3 logistic regression translated the causal findings into clinically interpretable effect sizes. By restricting models to the five strongest candidate domains per outcome, the resulting estimates focus on determinants with both predictive and causal support. For Mental Health, a one standard deviation increase in Stress was associated with 37% higher odds of cluster membership (AOR = 1.37), while Loneliness conferred 19% higher odds (AOR = 1.19). As visualized in Fig 3, these SDoH effects are comparable in magnitude to demographic factors such as female sex (AOR = 1.34), underscoring that social context carries weight alongside demographic structure in mental health risk.

For Cardiometabolic outcomes, age dominated the risk profile (AOR = 1.40 per standardized unit), as illustrated in Fig 4. Among SDoH domains, Stress (AOR = 1.14) and Neighborhood Disorder (AOR = 1.09) showed the strongest associations, consistent with evidence that neighborhood-level physical and social disorder contributes to cardiovascular risk through chronic stress exposure and reduced access to health-promoting resources [32]. The direction and relative ordering of these effects closely mirror the Stage 2 DML estimates, providing reassurance that the simpler parametric specification captures the same substantive relationships detected by the more flexible causal framework.

This concordance is noteworthy because the clinical utility of risk models depends on transparent, communicable effect estimates [49]. While the DML framework offers advantages for causal estimation in high-dimensional settings, odds ratios can be readily communicated to clinicians and policymakers. The logistic regression models achieved AUCs of 0.682 (Mental Health) and 0.630 (Cardiometabolic), retaining most of the XGBoost discriminative capacity (ΔAUC = 0.019 and 0.032, respectively). This interpretability–accuracy trade-off is acceptable for clinical applications where transparent risk communication is prioritized, and the retained AUC levels are comparable to those reported in scoping reviews of SDoH-enhanced prediction models [6, 7].

### 4.5 Heterogeneity Across Demographic Subgroups

We observed substantial heterogeneity in SDoH–outcome relationships across demographic strata, with 24 significant interactions after FDR correction (10 for Mental Health; 14 for Cardiometabolic). These findings extend the moderation patterns documented in our prior analysis of major depressive disorder [53] to a broader set of chronic disease outcomes, revealing that the differential vulnerability of demographic subgroups to social determinants is not limited to a single condition.

For Cardiometabolic outcomes, the effect of Stress was markedly stronger among Hispanic participants (AOR = 1.26 vs. 1.05 in the reference group, a 1.20-fold difference) and Black participants (1.19-fold). These amplified stress–cardiometabolic associations are consistent with the “weathering” hypothesis [40], which proposes that chronic exposure to social and economic adversity leads to accelerated physiological deterioration among racially minoritized populations. The fact that stress effects were amplified across all non-White racial/ethnic groups suggests that structurally patterned social stressors, rather than any single cultural factor, drive this differential vulnerability, aligning with fundamental-cause theory [16] and evidence that structural racism shapes health inequities through multiple reinforcing pathways [28].

Neighborhood Access showed differential effects for Cardiometabolic outcomes across multiple groups, with effects 1.10–1.11-fold stronger among Black, Hispanic, and Asian participants. This widespread amplification likely reflects structural disparities in neighborhood investment, where limited access to recreational facilities, healthy food sources, and healthcare disproportionately impacts marginalized communities [33]. Conversely, the weaker Neighborhood Disorder effects among Black participants for both Mental Health (0.96-fold) and Cardiometabolic (0.92-fold) outcomes merit careful interpretation. One possible explanation is psychological adaptation: individuals with greater lifetime exposure to neighborhood disorder may develop coping strategies that attenuate the health impact of these exposures, though such adaptation does not eliminate the underlying structural disadvantage.

Religion/Spirituality demonstrated stronger protective effects for Mental Health among Hispanic (0.94-fold), Asian (0.93-fold), and gender/sexual minority populations (0.95-fold), suggesting that religious and spiritual coping resources may be particularly impactful in communities where these institutions serve as primary sources of social support and cultural continuity [41, 54]. The enhanced protective effect among gender/sexual minority individuals is notable given the complex relationship between LGBTQIA2+ identity and religious institutions, and may reflect the protective role of affirming spiritual communities that buffer minority stress. In our prior analysis of major depressive disorder, social cohesion showed a paradoxical interaction with Asian race/ethnicity [53]; the current multi-indication framework reveals a broader pattern in which Religion/Spirituality serves as a protective factor whose strength varies across populations, warranting further investigation into the specific mechanisms driving these differential effects.

The Discrimination × Hispanic interaction for Mental Health (AOR = 1.06) aligns with evidence that experiences of discrimination carry particularly strong mental health consequences for Hispanic populations, potentially reflecting the compounding effects of anti-immigrant sentiment, language-based discrimination, and acculturative stress [20]. The attenuation of Loneliness effects among gender/sexual minority individuals (0.96-fold) may reflect the development of alternative social networks and community resilience within LGBTQIA2+ communities, though further research is needed to understand this pattern. These interaction patterns collectively reinforce the need for subgroup-aware interpretation when translating risk signals into clinical interventions and highlight the importance of culturally tailored approaches to SDoH-informed care.

### 4.6 Methodological Contributions

This study contributes methodologically by demonstrating a structured, multi-stage framework for integrating predictive machine learning, causal inference, and interpretable statistical modeling within a single analytical pipeline. As discussed in Section 1.3, relationships between SDoH and health outcomes are typically high-dimensional, nonlinear, and characterized by correlated exposures. Traditional regression models may struggle to identify relevant predictors under these conditions [44], while purely predictive machine learning models do not directly support causal interpretation. The staged approach implemented here addresses this tension by assigning distinct inferential roles to complementary methods.

In this framework, flexible machine learning models first identify important predictors and characterize complex relationships within the high-dimensional SDoH feature space. Causal estimation is then conducted separately using Double Machine Learning [48], which allows valid inference in the presence of high-dimensional confounding. Subsequent logistic regression translates these findings into clinically interpretable effect estimates, and formal interaction analysis assesses heterogeneity across demographic groups. By separating predictive discovery from causal estimation and interpretation, this design reduces the risk of conflating predictive importance with causal influence, a concern that is particularly salient in SDoH research where many domains are correlated with one another and with demographic characteristics.

More broadly, this staged design provides a practical template for researchers working with complex health datasets. The convergence of findings across stages strengthens confidence in the substantive conclusions beyond what any single analytical approach could provide. Stress, for example, emerges as the strongest predictor (Stage 1), the largest causal contributor (Stage 2), the most clinically impactful domain (Stage 3), and the most heterogeneous across subgroups (Stage 4).

### 4.7 Limitations

Results should be interpreted in light of several limitations.

#### Selection bias

The All of Us Research Program employs convenience sampling with voluntary enrollment, potentially selecting for individuals with greater health literacy, technology access, and healthcare engagement. The sample composition (69.6% White, 57.4% cisgender heterosexual female, 59.4% with college degree) may differ from the broader U.S. population. Additionally, 9.0% of participants had missing or unclassified race/ethnicity data, limiting the precision of subgroup analyses. Notably, the SDoH survey has only been available since November 2021 and is administered as an optional follow-up; individuals who did not participate in this survey were excluded from our analytical sample. This additional selection layer may introduce bias if SDoH survey completion is correlated with the social exposures under study.

#### Measurement

SDoH exposures were derived from self-reported surveys, introducing potential recall and social desirability bias. Survey-based assessment may inadequately capture objective social conditions that could be obtained through geocoded administrative data. The cross-sectional nature of SDoH measurement precludes assessment of exposure timing, duration, or developmental periods of vulnerability. Importantly, the lower internal consistency observed for Stress (*α* = 0.41) and Neighborhood Disorder (*α* = 0.58) suggests that these constructs may be multidimensional, and future research may benefit from exploring alternative operationalization strategies or subtyping approaches for these domains.

#### Outcome ascertainment

Health outcomes combined self-report and EHR extraction, each with distinct limitations. Self-reported conditions may reflect diagnostic access rather than true prevalence, a concern that is particularly relevant for mental health conditions where racial and ethnic disparities in healthcare utilization are well documented. As we noted in our prior work [53], the lower likelihood of mental health diagnosis among racial/ethnic minority groups may partly reflect disparities in mental healthcare access rather than genuine differences in disease burden. The clustering approach, while addressing co-occurrence, may also obscure heterogeneity within clusters depression and substance use disorder, for instance, may have distinct SDoH profiles despite grouping together.

#### Causal assumptions

DML estimates assume no unmeasured confounding, positivity, and consistency assumptions that cannot be fully verified in observational data. Residual confounding by unmeasured genetic, behavioral, or environmental factors may bias causal estimates. The cross-sectional design precludes establishing temporal precedence, leaving open the possibility of reverse causality, for example, individuals with depression may report higher stress levels as a consequence rather than a cause of their condition.

#### Social position

Our analysis models demographic characteristics and SDoH domains as largely independent predictors and moderators, yet the relationships between objective sociodemographic factors and subjective assessments of social position are complex. As the social position literature emphasizes, subjective social standing depends not only on observable characteristics but also on how individuals experience society and perceive their relative position [26]. The weaker-than-expected influence of some SDoH interactions may reflect limitations in capturing these nuanced dimensions of social position within the All of Us survey framework. Future research should expand beyond regression analysis to include structural equation models that encompass mediation and moderation effects of social position on SDoH and their interacting roles on health outcomes.

#### Multiple testing

Despite FDR correction for interaction analyses, the large number of tests conducted across four stages increases the risk of false positives. Replication in independent cohorts is essential before drawing strong conclusions about specific interactions.

### 4.8 Future Directions

Several directions could extend these findings. External validation in independent cohorts (e.g., UK Biobank, Million Veteran Program) would strengthen confidence in the robustness of SDoH health relationships. If All of Us accumulates longitudinal follow-up, future analyses should examine temporal relationships between SDoH exposures and incident outcomes using time-varying exposure models, which would address the current limitation of cross-sectional SDoH measurement and enable stronger causal claims about exposure timing and duration.

A critical translational step is evaluating whether condition-specific SDoH screening linked to referral pathways improves downstream outcomes through pragmatic trials. Integration of biomarker data available in All of Us could illuminate the biological mechanisms mediating SDoH health relationships, particularly the allostatic load pathways hypothesized to underlie the amplified stress effects observed among minoritized populations [40]. Structural equation models incorporating mediation through social position could further clarify the pathways through which demographic characteristics and SDoH jointly influence health outcomes.

A lifespan approach to social determinants of mental health is crucial, as improving early-life mental health conditions could have implications for population health across the life course. Effective interventions targeting early-life social exposures have the potential to reduce the risk of chronic disease in later life. Community-engaged research partnerships should develop and test culturally adapted interventions addressing the SDoH domains most impactful for specific populations, particularly stress management and discrimination-coping programs for Hispanic and Black communities, given the amplified stress and discrimination effects observed in these groups. Finally, alternative approaches to operationalizing SDoH subdomains from the All of Us measurements may capture dimensions of social context not fully addressed by our current domain structure.

### 4.9 Conclusions

Experiential social determinants of health, particularly stress, loneliness, and discrimination, emerge as the strongest and most consistent contributors to Mental Health risk, with effects that are robust across predictive, causal, and interpretable modeling stages. Cardiometabolic outcomes show smaller and more diffuse associations with social exposures, suggesting that these conditions reflect longer-term behavioral and physiological pathways where age and structural neighborhood characteristics play a more prominent role. The observed heterogeneity in SDoH effects across demographic groups, notably the amplified stress cardiometabolic association among Black and Hispanic participants and the differential protective role of Religion/Spirituality across racial/ethnic and sexual minority populations highlights that the health consequences of social exposures vary substantially across subpopulations.

Methodologically, the staged analytical pipeline combining machine learning for variable selection, double machine learning for causal estimation, and logistic regression for clinical interpretability demonstrates a practical framework for rigorously identifying and communicating actionable social risk factors. The convergence of results across these complementary approaches provides support for the notion that the domains identified represent meaningful causal pathways rather than purely predictive signals.

Taken together, these findings support condition-specific SDoH assessment rather than universal screening, allowing clinicians to focus on the social determinants most likely to influence the patient’s specific health risk. Integrating such insights into clinical practice requires validation in diverse populations and evaluation of targeted screening protocols. This work provides both substantive and methodological guidance for translating SDoH research into interventions that can effectively address social contributors to health disparities.

## Supporting Information

**S1 Appendix.** Technical Supplement: SDoH Categories and Constituent Questions.

**S2 Appendix.** Detailed Computational Results: TRIPOD+AI Checklist, Full SHAP Rankings, Full DML Results, Full Interaction Analysis.

**S3 Appendix.** Algorithm Pseudocode: Data Processing and Analytical Stages 1–4.

## Acknowledgments

We gratefully acknowledge the *All of Us* participants for their contributions, without whom this research would not have been possible. We also thank the National Institutes of Health’s *All of Us* Research Program for making available the participant data examined in this study. We also acknowledge Good Systems, a research grand challenge at The University of Texas at Austin that has been motivational for our team in our study of responsible artificial intelligence.

## Ethics Statement

This study was conducted using data from the *All of Us* Research Program. All participants provided informed consent for research use of their data as part of enrollment in the program. The *All of Us* Research Program is approved by the *All of Us* Institutional Review Board (IRB) at Vanderbilt University Medical Center. This specific analysis was conducted under the *All of Us* Registered Tier Data Access policies, which permit secondary analysis of de-identified data for approved research purposes.

## Data Availability Statement

The data underlying this study are available through the *All of Us* Research Program Researcher Workbench. Due to participant privacy protections and the *All of Us* Data Use Agreement, data cannot be shared publicly. Qualified researchers may apply for access to the *All of Us* Researcher Workbench at https://www.researchallofus.org. The analytical code used in this study is available upon reasonable request to the corresponding author and will be deposited in a public repository upon acceptance for publication.

## Competing Interests

The authors have declared that no competing interests exist.

## Author Contributions

**Conceptualization:** Matt Kammer-Kerwick, Yug Dave. **Data curation:** Yug Dave. **Formal analysis:** Yug Dave, Matt Kammer-Kerwick, Vidhi Parekh, Luke McDonald. **Funding acquisition:** Matt Kammer-Kerwick, S. Craig Watkins. **Investigation:** Yug Dave, Matt Kammer-Kerwick. **Methodology:** Matt Kammer-Kerwick, Yug Dave. **Project administration:** Matt Kammer-Kerwick, S. Craig Watkins. **Resources:** Matt Kammer-Kerwick, S. Craig Watkins. **Software:** Yug Dave, Matt Kammer-Kerwick. **Supervision:** Matt Kammer-Kerwick. **Validation:** Yug Dave, Matt Kammer-Kerwick. **Visualization:** Yug Dave, Vidhi Parekh, Luke McDonald. **Writing – original draft:** Matt Kammer-Kerwick, Yug Dave, Vidhi Parekh, Luke McDonald. **Writing – review & editing:** Matt Kammer- Kerwick, Yug Dave, Vidhi Parekh, Luke McDonald.

## S1 Text: Supporting Information

### Box A in S1 Text — SDoH Categories and Constituent Questions

The 12 SDoH domain features were constructed from standardized *All of Us* survey items grouped into predefined domains using established domain mappings [1]. Rather than using principal component analysis, we summarized each domain using a SHAP-weighted aggregation approach: global item importance weights were derived from mean absolute SHAP values computed from the Combined XGBoost model, and each domain score was computed as a weighted average of its constituent standardized items [2, 3]. The categories and their corresponding `question_concept_id`s from the *All of Us* dataset are detailed in Table 1.

**Table 1: Table A in S1 Text.**
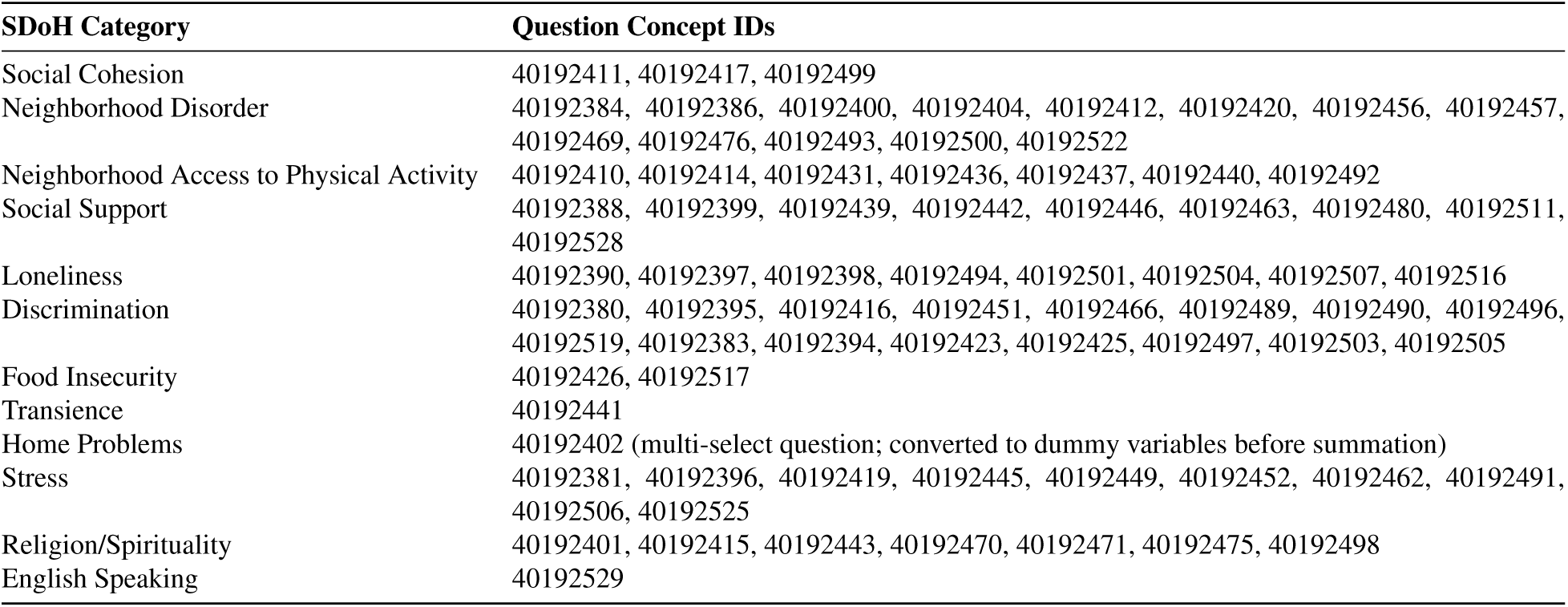
Social Determinant of Health (SDoH) Categories and Source Question IDs.

### Box B in S1 Text — Detailed Computational Results

#### TRIPOD+AI Checklist

The Transparent Reporting of a multivariable prediction model for Individual Prognosis Or Diagnosis with Artificial Intelligence (TRIPOD+AI) checklist is provided as a separate Supporting Information file. This checklist documents adherence to recommended reporting guidelines for prediction models that incorporate machine learning methods [4].

#### Complete Domain-Level SHAP Importance Rankings

Table 2 presents the complete domain-level SHAP importance values for all SDoH and demographic features across both outcome clusters. Values represent the sum of mean absolute SHAP values across all items within each domain. Higher values indicate greater average contribution to the model’s predicted probability of the outcome.

**Table 2: Table B in S1 Text.** Complete Domain-Level SHAP Importance by Outcome. Values represent total SHAP importance (sum of mean |SHAP| across items in domain). Top 3 domains per outcome are marked with †.

| Domain | Mental Health | Cardiometabolic |
| --- | --- | --- |
| *SDoH Domains* |  |  |
| Stress | 0.361<sup>†</sup> | 0.141 |
| Discrimination | 0.283<sup>†</sup> | 0.225<sup>†</sup> |
| Religion/Spirituality | 0.272<sup>†</sup> | 0.204 |
| Loneliness | 0.254 | 0.108 |
| Neighborhood Disorder | 0.179 | 0.232<sup>†</sup> |
| Social Support | 0.144 | 0.095 |
| Neighborhood Access | 0.119 | 0.174 |
| Social Cohesion | 0.039 | 0.032 |
| English Speaking | 0.032 | 0.004 |
| Food Insecurity | 0.017 | 0.049 |
| Transience | 0.012 | 0.008 |
| Home Problems | 0.010 | 0.006 |
| *Demographic Features* |  |  |
| Age | 0.130 | 0.284<sup>†</sup> |
| Female (Cis-Het) | 0.189 | 0.084 |
| Gender/Sexual Minority | 0.089 | 0.031 |
| Black | 0.082 | 0.027 |
| Asian | 0.059 | 0.016 |
| Hispanic | 0.051 | 0.023 |
| Household Income | 0.050 | 0.074 |
| Education Level | 0.038 | 0.097 |
| Other Race/Ethnicity | 0.009 | 0.001 |
| Housing Insecurity | 0.009 | 0.006 |

#### Domain Review Summary Tables

Tables 3 and 4 present the domain-level evidence summaries used to determine which SDoH domains were carried forward into Stages 3 and 4. Each domain was evaluated using four criteria: SHAP importance rank from Stage 1, the DML average treatment effect (ATE) magnitude and statistical significance from Stage 2, the variance inflation factor (VIF) from the domain correlation matrix, and Cronbach’s alpha (*α*) for internal consistency. Domains were classified as *strong_candidate*, *review_carefully*, or *weak_candidate* based on converging evidence across these criteria. Only *strong_candidate* domains were retained for downstream clinical modeling.

**Table 3: Table C in S1 Text.** Domain Review Summary: Mental Health Outcome. Domains ranked by SHAP importance from Stage 1 with supporting evidence from Stage 2 DML, multicollinearity diagnostics (VIF), and internal consistency (*α*). Only the five *strong_candidate* domains were carried forward into Stages 3 and 4.

| Domain | SHAP Rk | DML ATE | DML $p$ | VIF | $\alpha$ | Label |
| --- | --- | --- | --- | --- | --- | --- |
| Stress | 1 | 0.0931 | <0.001 | 1.5 | 0.41 | strong_candidate |
| Discrimination | 2 | 0.0635 | <0.001 | 1.6 | 0.93 | strong_candidate |
| Religion/Spirituality | 3 | -0.0219 | <0.001 | 1.1 | 0.93 | strong_candidate |
| Loneliness | 4 | 0.0769 | <0.001 | 1.6 | 0.67 | strong_candidate |
| Neighborhood Disorder | 5 | 0.0285 | <0.001 | 1.3 | 0.58 | strong_candidate |
| Social Support | 6 | 0.0005 | 0.582 | 1.3 | 0.93 | review_carefully |
| Neighborhood Access | 7 | 0.0119 | <0.001 | 1.2 | 0.74 | review_carefully |
| Social Cohesion | 8 | 0.0011 | 0.270 | 1.2 | 0.84 | review_carefully |
| English Speaking | 9 | 0.0356 | <0.001 | 1.1 | N/A | review_carefully |
| Food Insecurity | 10 | 0.0201 | <0.001 | 1.3 | 0.90 | review_carefully |
| Transience | 11 | -0.0014 | 0.179 | 1.1 | N/A | weak_candidate |
| Home Problems | 12 | 0.0165 | <0.001 | 1.1 | N/A | review_carefully |

**Table 4: Table D in S1 Text.** Domain Review Summary: Cardiometabolic Outcome. Domains ranked by SHAP importance from Stage 1 with supporting evidence from Stage 2 DML, multicollinearity diagnostics (VIF), and internal consistency (*α*). Only the five *strong_candidate* domains were carried forward into Stages 3 and 4.

| Domain | SHAP Rk | DML ATE | DML $p$ | VIF | $\alpha$ | Label |
| --- | --- | --- | --- | --- | --- | --- |
| Neighborhood Disorder | 1 | 0.0255 | <0.001 | 1.3 | 0.58 | strong_candidate |
| Discrimination | 2 | 0.0227 | <0.001 | 1.5 | 0.93 | strong_candidate |
| Religion/Spirituality | 3 | 0.0054 | <0.001 | 1.0 | 0.93 | strong_candidate |
| Neighborhood Access | 4 | 0.0112 | <0.001 | 1.2 | 0.74 | strong_candidate |
| Stress | 5 | 0.0357 | <0.001 | 1.5 | 0.41 | strong_candidate |
| Loneliness | 6 | 0.0231 | <0.001 | 1.3 | 0.67 | review_carefully |
| Social Support | 7 | 0.0245 | <0.001 | 1.3 | 0.93 | review_carefully |
| Food Insecurity | 8 | 0.0004 | 0.674 | 1.3 | 0.90 | review_carefully |
| Social Cohesion | 9 | 0.0237 | <0.001 | 1.2 | 0.84 | review_carefully |
| Transience | 10 | -0.0073 | <0.001 | 1.1 | N/A | review_carefully |
| Home Problems | 11 | 0.0017 | 0.059 | 1.1 | N/A | review_carefully |
| English Speaking | 12 | 0.0315 | <0.001 | 1.1 | N/A | review_carefully |

#### Full DML Results with Heterogeneous Treatment Effects

Table 5 presents the complete Average Treatment Effect (ATE) estimates from Double Machine Learning for all twelve SDoH domains across both outcome clusters. Conditional ATEs within demographic strata are visualized in Fig 3.

**Table 5: Table E in S1 Text.** Full DML Average Treatment Effects Across All SDoH Domains. Values represent the estimated change in outcome probability per 1-SD increase in SDoH domain exposure, with 95% confidence intervals.

| SDoH Domain | Mental Health | Cardiometabolic |
| --- | --- | --- |
| Stress | 0.0931*** (0.0910–0.0949) | 0.0357*** (0.0339–0.0375) |
| Loneliness | 0.0769*** (0.0750–0.0788) | 0.0231*** (0.0212–0.0250) |
| Discrimination | 0.0635*** (0.0615–0.0655) | 0.0227*** (0.0207–0.0244) |
| English Speaking | 0.0356*** (0.0339–0.0376) | 0.0315*** (0.0296–0.0335) |
| Neighborhood Disorder | 0.0285*** (0.0269–0.0305) | 0.0255*** (0.0239–0.0271) |
| Food Insecurity | 0.0201*** (0.0179–0.0223) | 0.0004 (–0.0017–0.0024) |
| Home Problems | 0.0165*** (0.0142–0.0185) | 0.0017 (–0.0002–0.0033) |
| Neighborhood Access | 0.0119*** (0.0099–0.0137) | 0.0112*** (0.0097–0.0128) |
| Social Cohesion | 0.0011 (–0.0007–0.0029) | 0.0237*** (0.0219–0.0258) |
| Social Support | 0.0005 (–0.0013–0.0024) | 0.0245*** (0.0233–0.0261) |
| Religion/Spirituality | –0.0219*** (–0.0239––0.0202) | 0.0054*** (0.0040–0.0070) |
| Transience | –0.0014 (–0.0034–0.0005) | –0.0073*** (–0.0089––0.0056) |

\* $p < 0.05$ ; \*\* $p < 0.01$ ; \*\*\* $p < 0.001$

#### Full Interaction Analysis Results

Table 6 presents the complete set of significant SDoH × demographic interaction terms from Stage 4, including all 24 interactions that survived Benjamini–Hochberg FDR correction at *q <* 0.05 (10 for Mental Health; 14 for Cardiometabolic).

**Table 6: Table F in S1 Text.** Complete Significant SDoH × Demographic Interactions. Interaction AORs represent the multiplicative modifier applied to the reference-group main effect for the indicated subgroup. All *p*-values are FDR-adjusted.

| Outcome | Interaction Term | AOR | 95% CI | $p$-value |
| --- | --- | --- | --- | --- |
| *Mental Health (10 significant interactions)* |  |  |  |  |
| Mental Health | Religion/Spirituality × Hispanic | 0.94 | 0.91–0.96 | <0.0001*** |
| Mental Health | Stress × Asian | 1.13 | 1.05–1.20 | 0.0005*** |
| Mental Health | Religion/Spirituality × Sexual Minority | 0.95 | 0.93–0.98 | 0.0007*** |
| Mental Health | Discrimination × Hispanic | 1.06 | 1.02–1.09 | 0.0019** |
| Mental Health | Loneliness × Sexual Minority | 0.96 | 0.93–0.98 | 0.0024** |
| Mental Health | Neighborhood Disorder × Black | 0.96 | 0.93–0.99 | 0.0058** |
| Mental Health | Religion/Spirituality × Asian | 0.93 | 0.88–0.98 | 0.0097** |
| Mental Health | Neighborhood Disorder × Hispanic | 0.97 | 0.94–1.00 | 0.0327* |
| Mental Health | Religion/Spirituality × Other | 0.93 | 0.87–1.00 | 0.0390* |
| Mental Health | Neighborhood Disorder × Other | 0.93 | 0.87–1.00 | 0.0417* |
| *Cardiometabolic (14 significant interactions)* |  |  |  |  |
| Cardiometabolic | Stress × Hispanic | 1.20 | 1.17–1.24 | <0.0001*** |
| Cardiometabolic | Stress × Black | 1.19 | 1.15–1.23 | <0.0001*** |
| Cardiometabolic | Neighborhood Access × Black | 1.11 | 1.07–1.15 | <0.0001*** |
| Cardiometabolic | Neighborhood Access × Hispanic | 1.10 | 1.06–1.14 | <0.0001*** |
| Cardiometabolic | Neighborhood Disorder × Black | 0.92 | 0.90–0.95 | <0.0001*** |
| Cardiometabolic | Religion/Spirituality × Black | 1.08 | 1.04–1.12 | <0.0001*** |
| Cardiometabolic | Stress × Asian | 1.12 | 1.06–1.19 | <0.0001*** |
| Cardiometabolic | Stress × Other | 1.18 | 1.09–1.28 | <0.0001*** |
| Cardiometabolic | Neighborhood Access × Sexual Minority | 1.07 | 1.03–1.10 | <0.0001*** |
| Cardiometabolic | Discrimination × Black | 1.06 | 1.03–1.10 | 0.0002*** |
| Cardiometabolic | Neighborhood Access × Asian | 1.10 | 1.03–1.16 | 0.0030** |
| Cardiometabolic | Neighborhood Disorder × Hispanic | 0.96 | 0.93–0.99 | 0.0060** |
| Cardiometabolic | Discrimination × Hispanic | 1.04 | 1.01–1.08 | 0.0192* |
| Cardiometabolic | Neighborhood Disorder × Other | 0.92 | 0.85–0.99 | 0.0327* |

\* $p < 0.05$ ; \*\* $p < 0.01$ ; \*\*\* $p < 0.001$ (FDR-adjusted)

#### Internal Consistency of SDoH Domain Scores

**Table 7: Table G in S1 Text.** Internal Consistency (Cronbach’s Alpha) of SDoH Domain Scores. Cronbach’s alpha measures the internal consistency of a set of survey items intended to capture a common latent construct. Values above approximately 0.70 indicate acceptable reliability. Single-item domains do not permit alpha computation.

| Domain | N Items | Cronbach's $\alpha$ |
| --- | --- | --- |
| Social Cohesion | 3 | 0.84 |
| Neighborhood Disorder | 13 | 0.58 |
| Neighborhood Access | 7 | 0.74 |
| Social Support | 9 | 0.93 |
| Loneliness | 8 | 0.67 |
| Discrimination | 16 | 0.93 |
| Food Insecurity | 2 | 0.90 |
| Transience | 1 | N/A |
| Home Problems | 1 | N/A |
| Stress | 10 | 0.41 |
| Religion/Spirituality | 7 | 0.93 |
| English Speaking | 1 | N/A |

#### Stage 3 Logistic Regression Results

**Table 8: Table H in S1 Text.** Stage 3 Adjusted Odds Ratios — Mental Health. Model includes five strong-candidate SDoH domains and seven demographic covariates (12 predictors total). AORs represent the multiplicative change in odds per 1-SD increase in standardized domain score.

| Domain | AOR | 95% CI | $\alpha$ |
| --- | --- | --- | --- |
| Stress | 1.37*** | (1.36–1.39) | 0.41 |
| Discrimination | 1.11*** | (1.10–1.12) | 0.93 |
| Religion/Spirituality | 0.93*** | (0.93–0.94) | 0.93 |
| Loneliness | 1.19*** | (1.18–1.20) | 0.67 |
| Neighborhood Disorder | 1.03*** | (1.02–1.04) | 0.58 |
| Demo: Female (Cis-Het) | 1.34*** | (1.33–1.35) | — |
| Demo: Age | 1.06*** | (1.05–1.07) | — |
| Demo: Gender/Sexual Minority | 1.27*** | (1.25–1.28) | — |
| Demo: Black | 0.84*** | (0.83–0.84) | — |
| Demo: Asian | 0.81*** | (0.80–0.82) | — |
| Demo: Hispanic | 0.84*** | (0.84–0.85) | — |
| Demo: Household Income | 1.09*** | (1.08–1.09) | — |
| Model AUC: 0.682 \| XGBoost AUC: 0.701 \| ΔAUC: 0.019 | | | |

\* $p < 0.05$ ; \*\* $p < 0.01$ ; \*\*\* $p < 0.001$

**Table 9: Table I in S1 Text.** Stage 3 Adjusted Odds Ratios — Cardiometabolic. Model includes five strong-candidate SDoH domains and six demographic covariates (11 predictors total). AORs represent the multiplicative change in odds per 1-SD increase in standardized domain score.

| Domain | AOR | 95% CI | $\alpha$ |
| --- | --- | --- | --- |
| Neighborhood Disorder | 1.09*** | (1.08–1.10) | 0.58 |
| Discrimination | 1.02*** | (1.01–1.03) | 0.93 |
| Religion/Spirituality | 1.03*** | (1.02–1.04) | 0.93 |
| Neighborhood Access | 0.99* | (0.98–1.00) | 0.74 |
| Stress | 1.14*** | (1.13–1.15) | 0.41 |
| Demo: Age | 1.40*** | (1.38–1.41) | — |
| Demo: Education Level | 1.18*** | (1.17–1.19) | — |
| Demo: Female (Cis-Het) | 1.13*** | (1.12–1.14) | — |
| Demo: Household Income | 1.17*** | (1.16–1.18) | — |
| Demo: Gender/Sexual Minority | 1.06*** | (1.05–1.07) | — |
| Demo: Black | 0.92*** | (0.91–0.92) | — |
| Model AUC: 0.630 \| XGBoost AUC: 0.662 \| ΔAUC: 0.032 | | | |

\* $p < 0.05$ ; \*\* $p < 0.01$ ; \*\*\* $p < 0.001$

#### Stage 4 Reference-Group Baselines and Stratified Effects

**Table 10: Table J in S1 Text.** Stage 4 Main Effects (Reference-Group Baselines) from Interaction Models. AORs represent the SDoH domain effect for the reference group (White, cisgender heterosexual participants) per 1- SD increase in standardized domain score.

| SDoH Domain | Mental Health<br>AOR [95% CI] | Cardiometabolic<br>AOR [95% CI] |
| --- | --- | --- |
| Stress | 1.41 (1.39–1.42)*** | 1.05 (1.03–1.06)*** |
| Discrimination | 1.10 (1.09–1.12)*** | 0.95 (0.94–0.97)*** |
| Religion/Spirituality | 0.96 (0.95–0.97)*** | 1.05 (1.03–1.06)*** |
| Loneliness | 1.18 (1.16–1.19)*** | — |
| Neighborhood Disorder | 1.02 (1.01–1.03)*** | 1.07 (1.05–1.08)*** |
| Neighborhood Access | — | 0.99 (0.98–1.00) |
| **Model AUC** | **0.669** | **0.566** |

\* $p < 0.05$ ; \*\* $p < 0.01$ ; \*\*\* $p < 0.001$
“—” indicates domain not retained for that outcome.

**Table 11: Table K in S1 Text.** Stratified Effects for Significant Interactions. AORs are shown separately for the reference group (modifier = 0) and modifier group (modifier = 1). The reference-group AOR corresponds to the main effect from the interaction model; the modifier-group AOR is the product of the main effect and the interaction AOR.

| Interaction | Outcome | AOR (Ref) | AOR (Mod) | Ratio |
| --- | --- | --- | --- | --- |
| *Mental Health* |  |  |  |  |
| Religion/Spirituality × Hispanic | Mental Health | 0.96 | 0.90 | 0.94× weaker |
| Stress × Asian | Mental Health | 1.41 | 1.59 | 1.13× stronger |
| Religion/Spirituality × Sexual Minority | Mental Health | 0.96 | 0.92 | 0.95× weaker |
| Discrimination × Hispanic | Mental Health | 1.10 | 1.16 | 1.06× stronger |
| Loneliness × Sexual Minority | Mental Health | 1.18 | 1.13 | 0.96× weaker |
| *Cardiometabolic* |  |  |  |  |
| Stress × Hispanic | Cardiometabolic | 1.05 | 1.26 | 1.20× stronger |
| Stress × Black | Cardiometabolic | 1.05 | 1.24 | 1.19× stronger |
| Neighborhood Access × Black | Cardiometabolic | 0.99 | 1.10 | 1.11× stronger |
| Neighborhood Access × Hispanic | Cardiometabolic | 0.99 | 1.09 | 1.10× stronger |
| Neighborhood Disorder × Black | Cardiometabolic | 1.07 | 0.99 | 0.92× weaker |

### Box C in S1 Text — Algorithm Pseudocode

This section provides pseudocode for the full analysis pipeline, including data processing and feature engineering, followed by the Stage 1–Stage 4 estimation and inference procedures.

#### Algorithm 1 Data Processing and Feature Engineering Pipeline

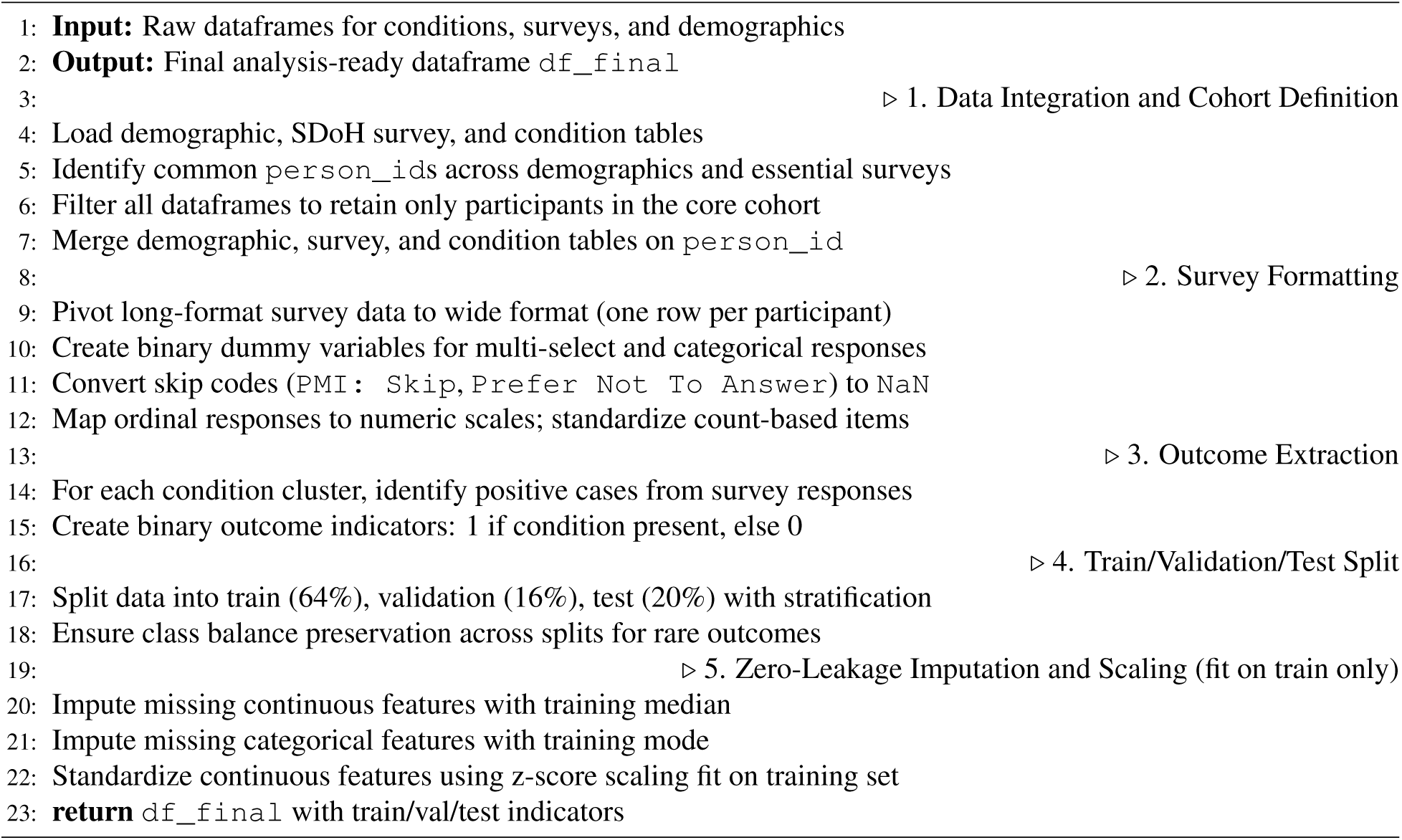

#### Algorithm 2 Stage 1: Discovery Analysis Pipeline

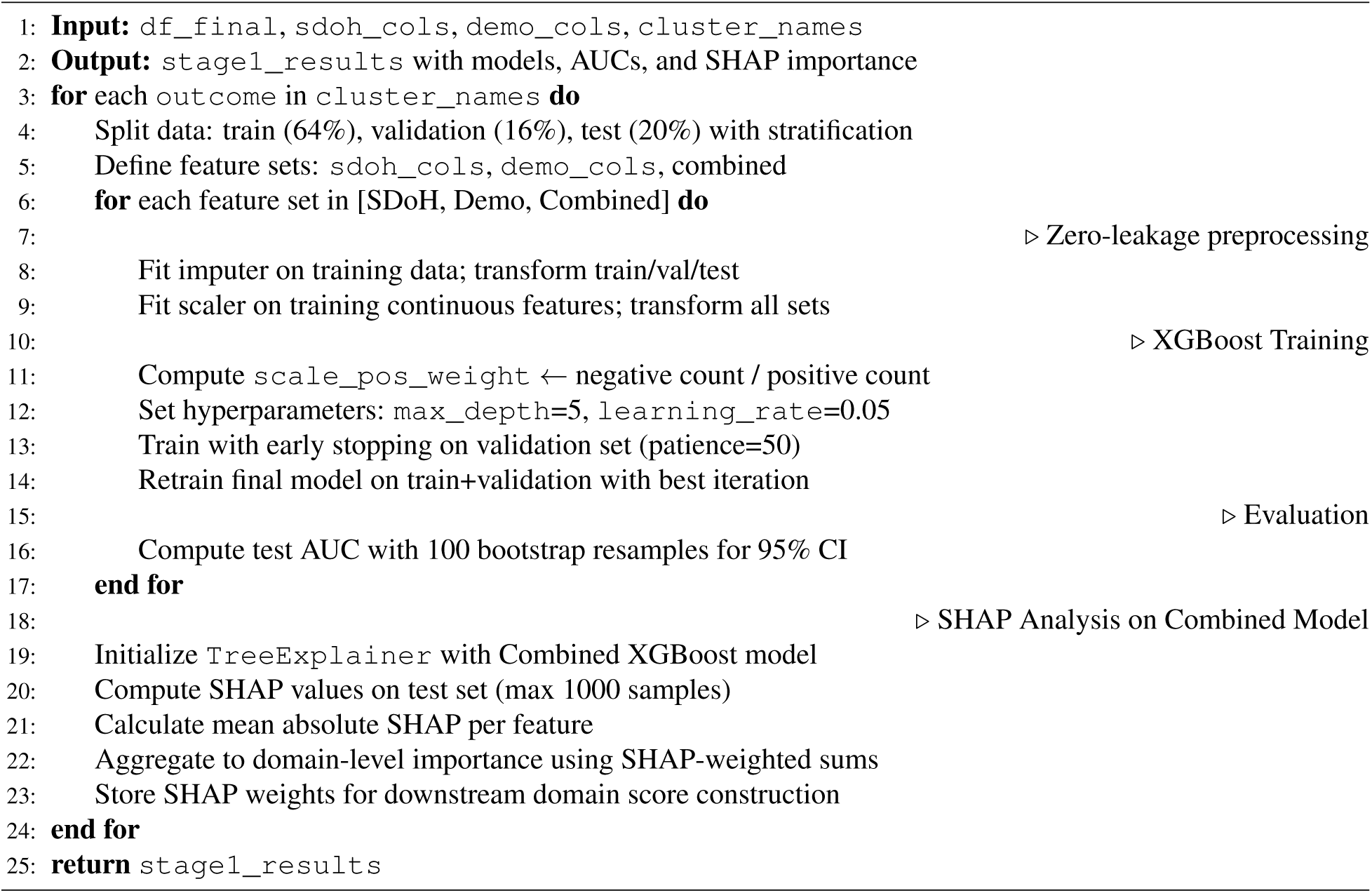

#### Algorithm 3 Stage 2: Double Machine Learning for Causal Effect Estimation

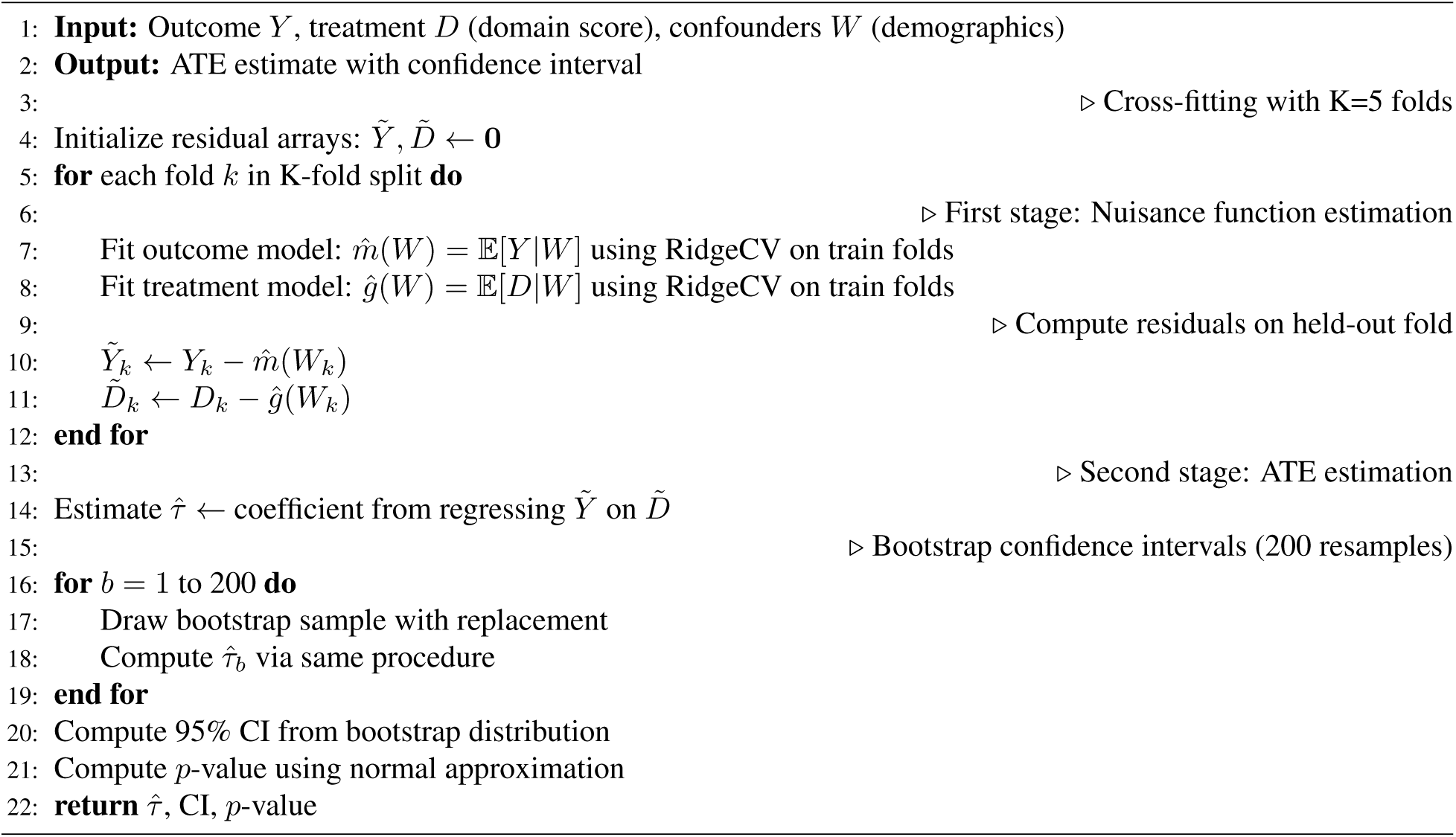

#### Algorithm 4 Stage 3: Logistic Regression for Clinical Interpretation

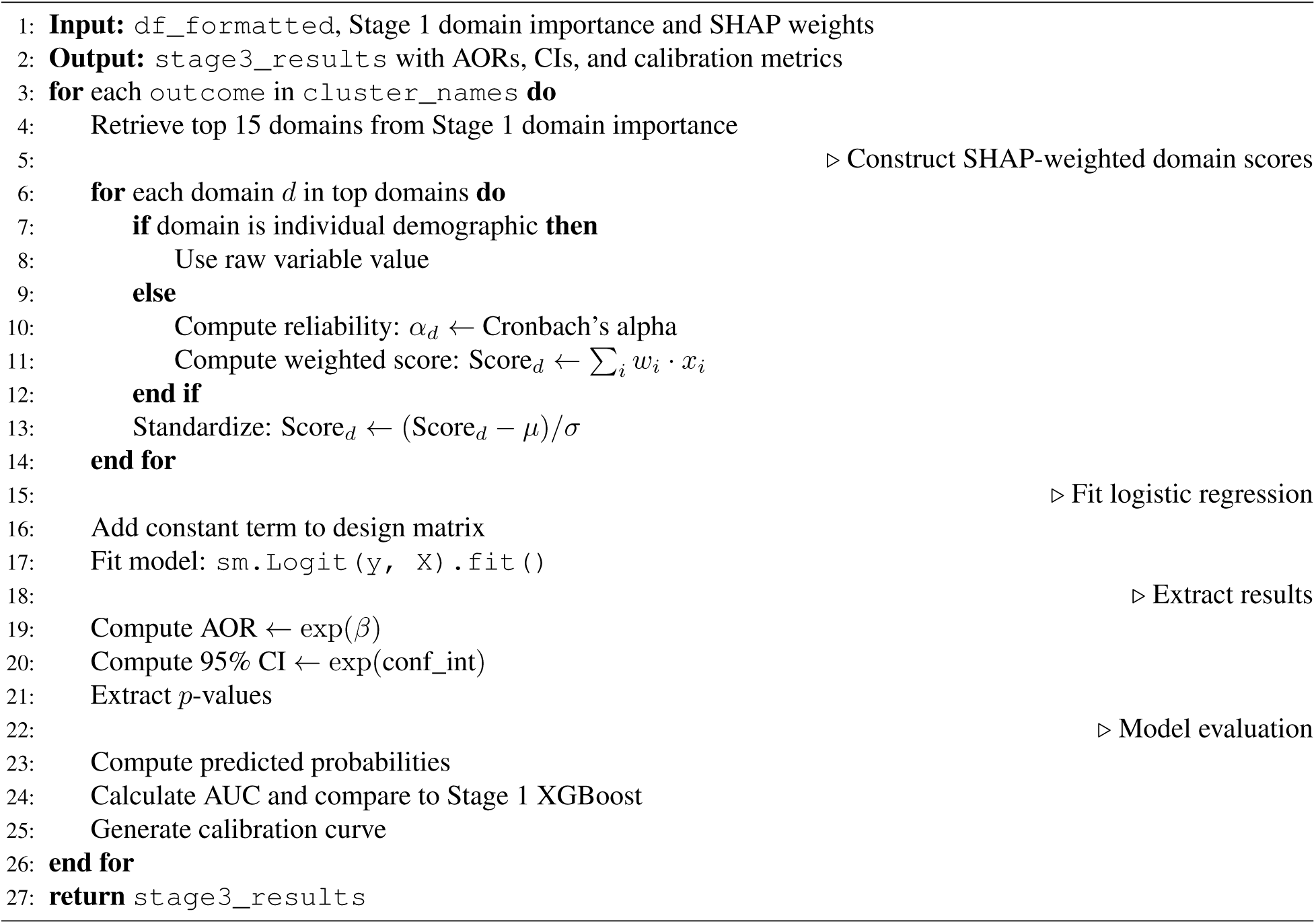

#### Algorithm 5 Stage 4: SDoH × Demographic Interaction Analysis

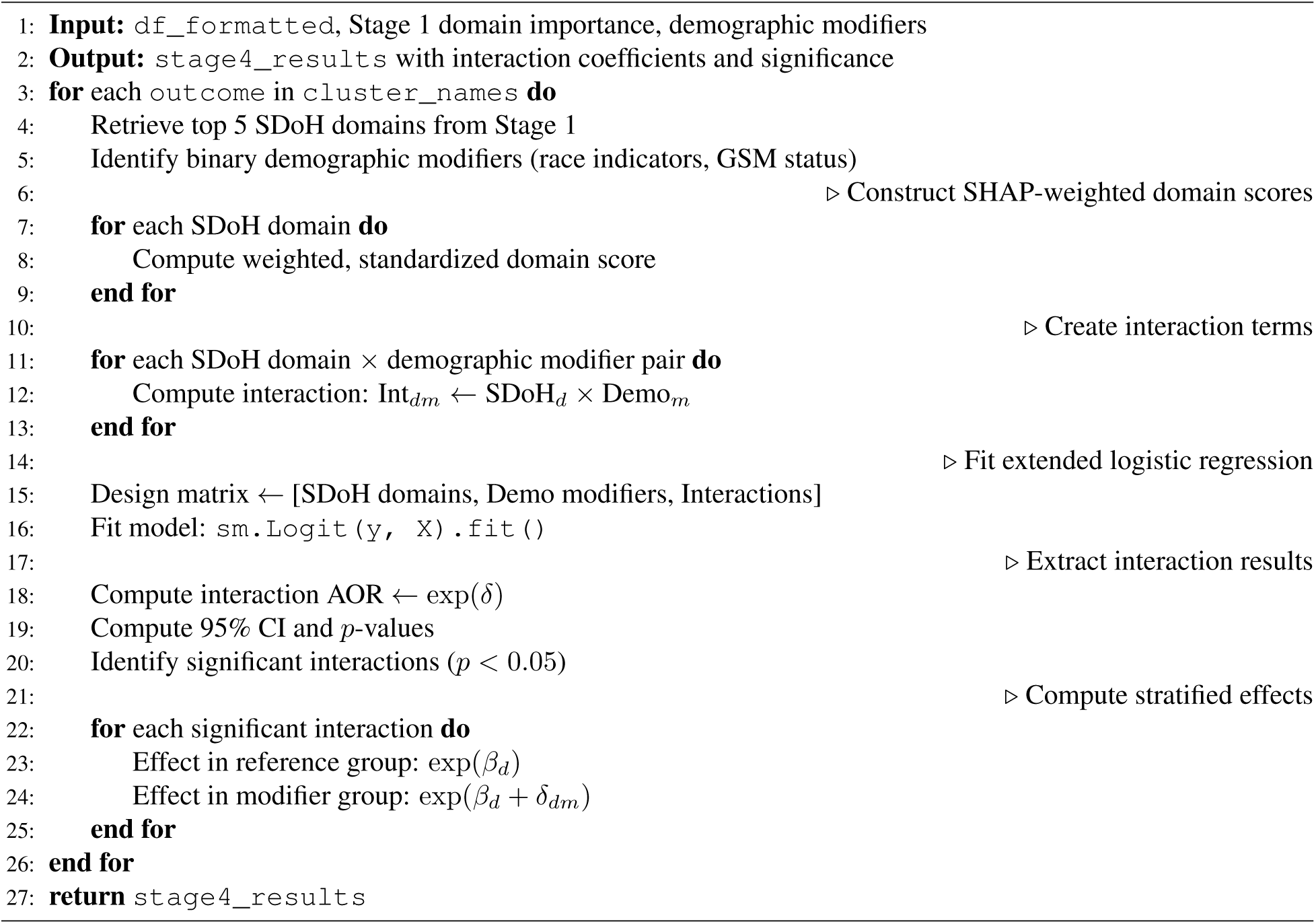

### Box D in S1 Text — Supplemental Figures

**Fig 1: Fig A in S1 Text.**
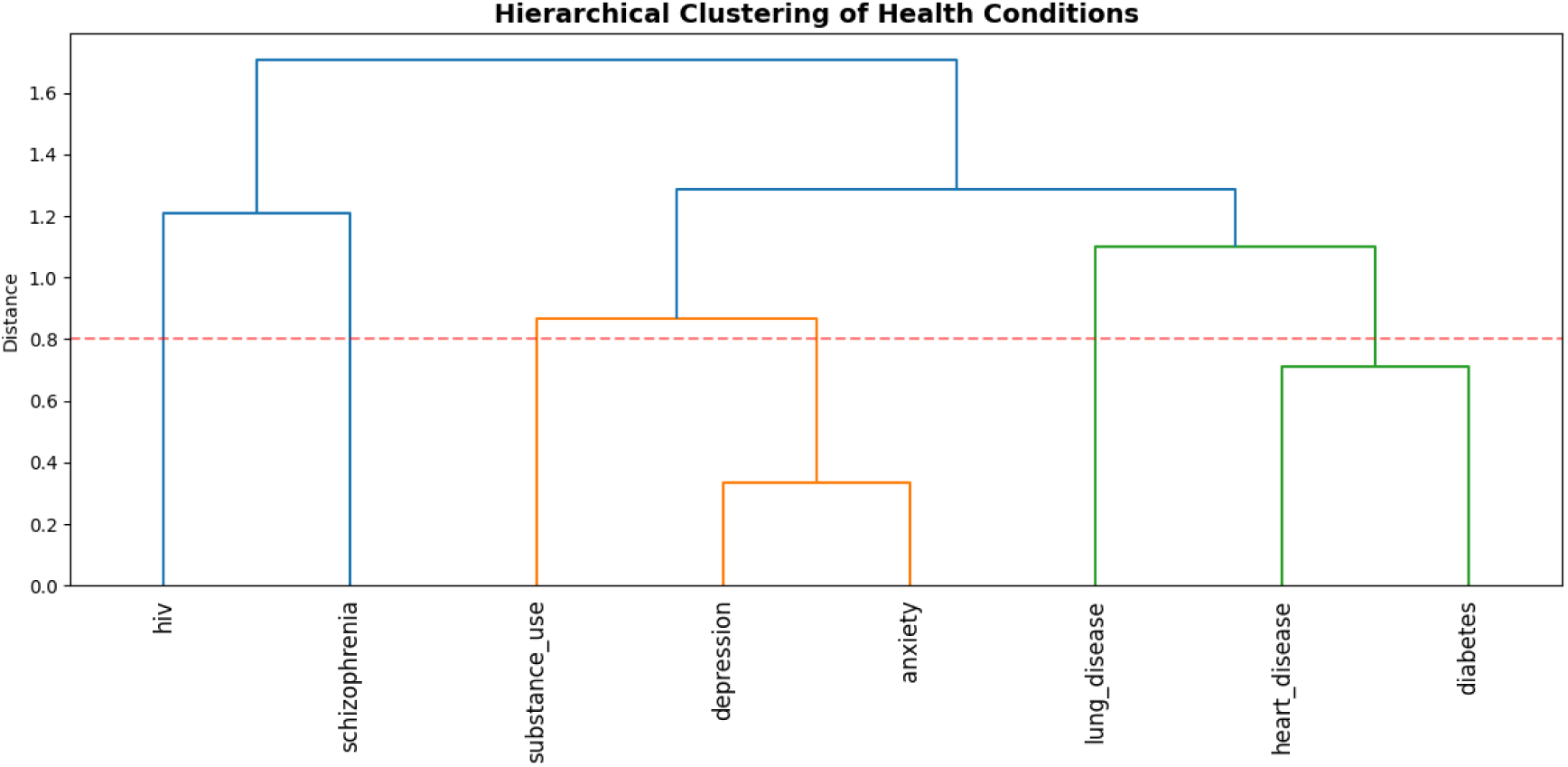
Hierarchical Clustering of Health Conditions. Dendrogram showing the clustering structure of eight health conditions based on co-occurrence patterns. The horizontal dashed line indicates the cut-off used for cluster assignment. Four clusters emerged: Mental Health (depression, anxiety, substance use disorder), Cardiometabolic (heart disease, diabetes, lung disease), Schizophrenia, and HIV.

**Fig 2: Fig B in S1 Text.**
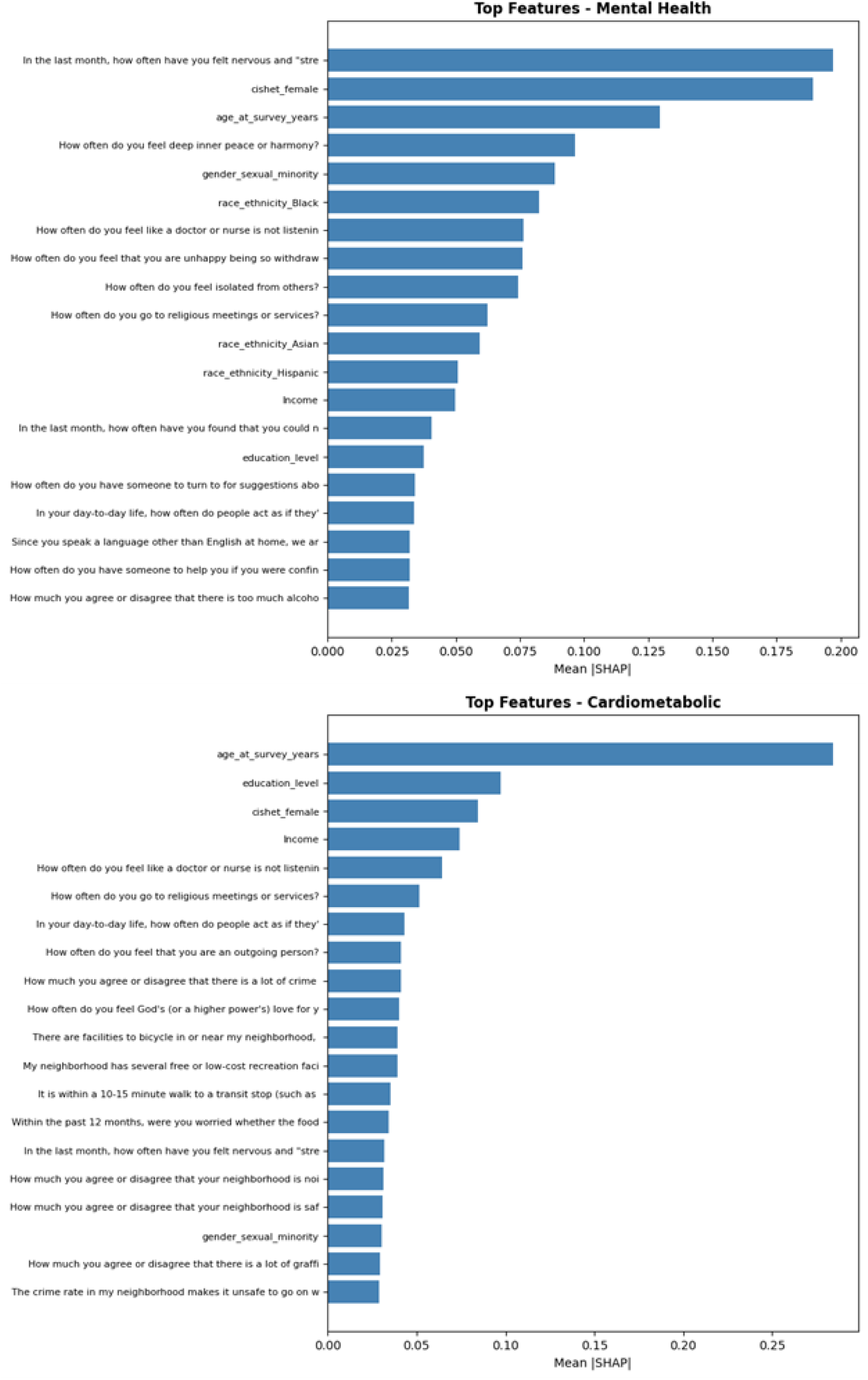
SHAP Feature Importance for Combined Models. Top 20 features ranked by mean absolute SHAP value for Mental Health (top) and Cardiom_5_e_5_tabolic (bottom) outcomes. Higher values indicate greater contribution to model predictions.

**Fig 3: Fig C in S1 Text.**
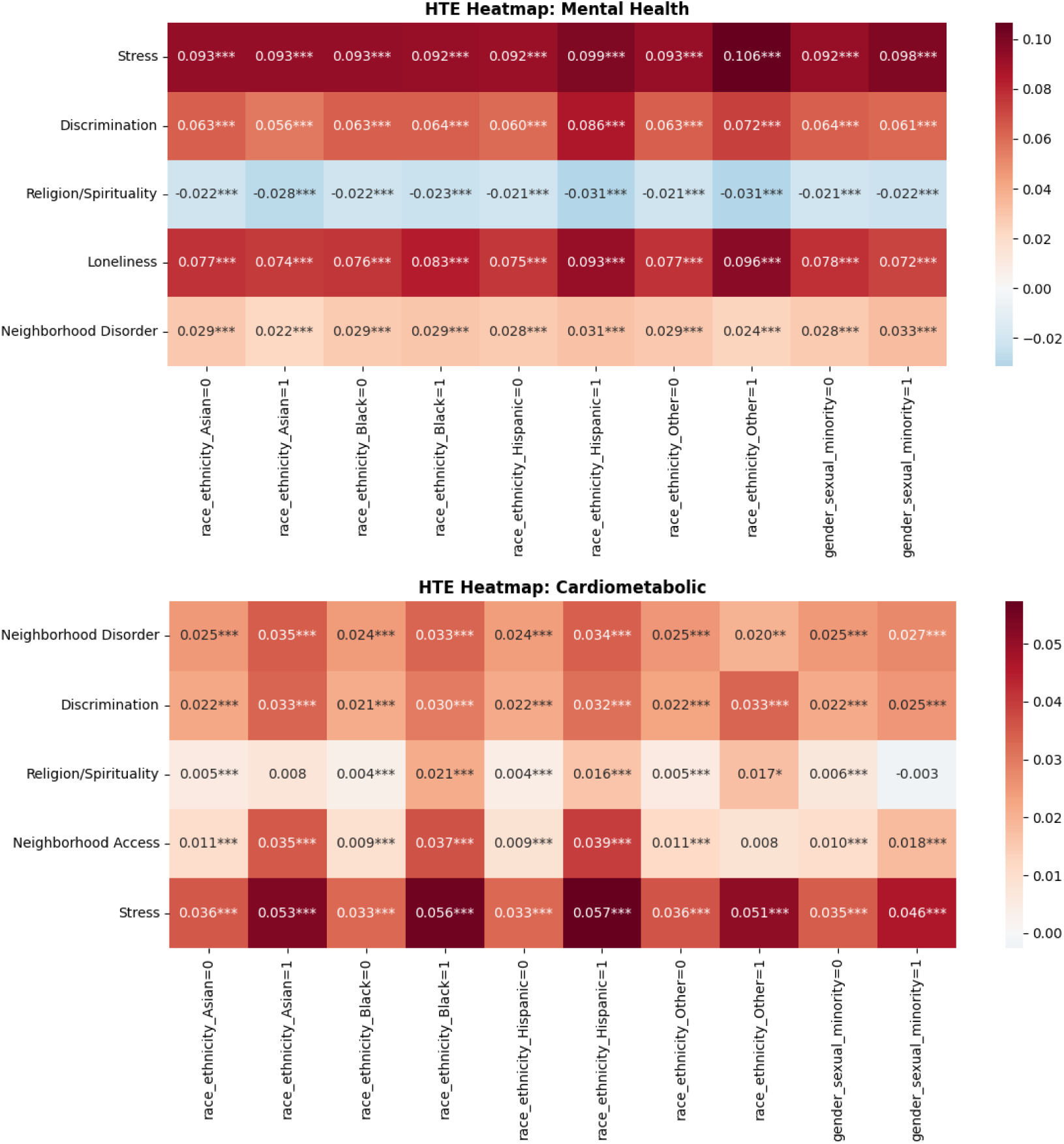
Heterogeneous Treatment Effect Heatmaps. Estimated ATEs across SDoH domains (rows) and demographic subgroups (columns) for Mental Health (top) and Cardiometabolic (bottom) outcomes. Asterisks indicate significance levels (* p < 0.05, ** p < 0.01, *** p < 0.001).

**Fig 4: Fig D in S1 Text.**
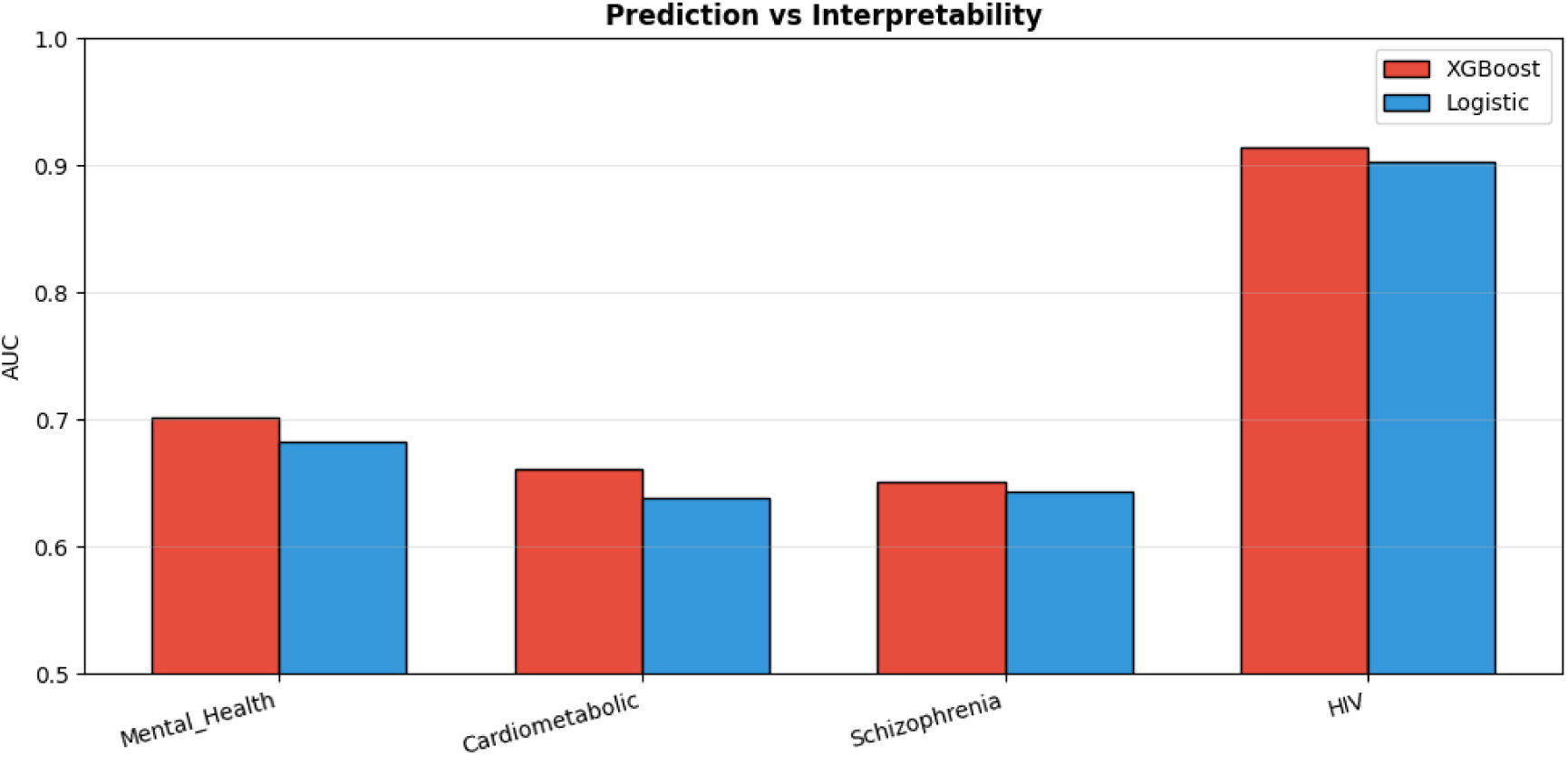
Prediction vs. Interpretability Trade-off. Comparison of predictive performance between XGBoost (Stage 1) and interpretable Logistic Regression (Stage 3) models across outcome clusters.

**Fig 5: Fig E in S1 Text.**
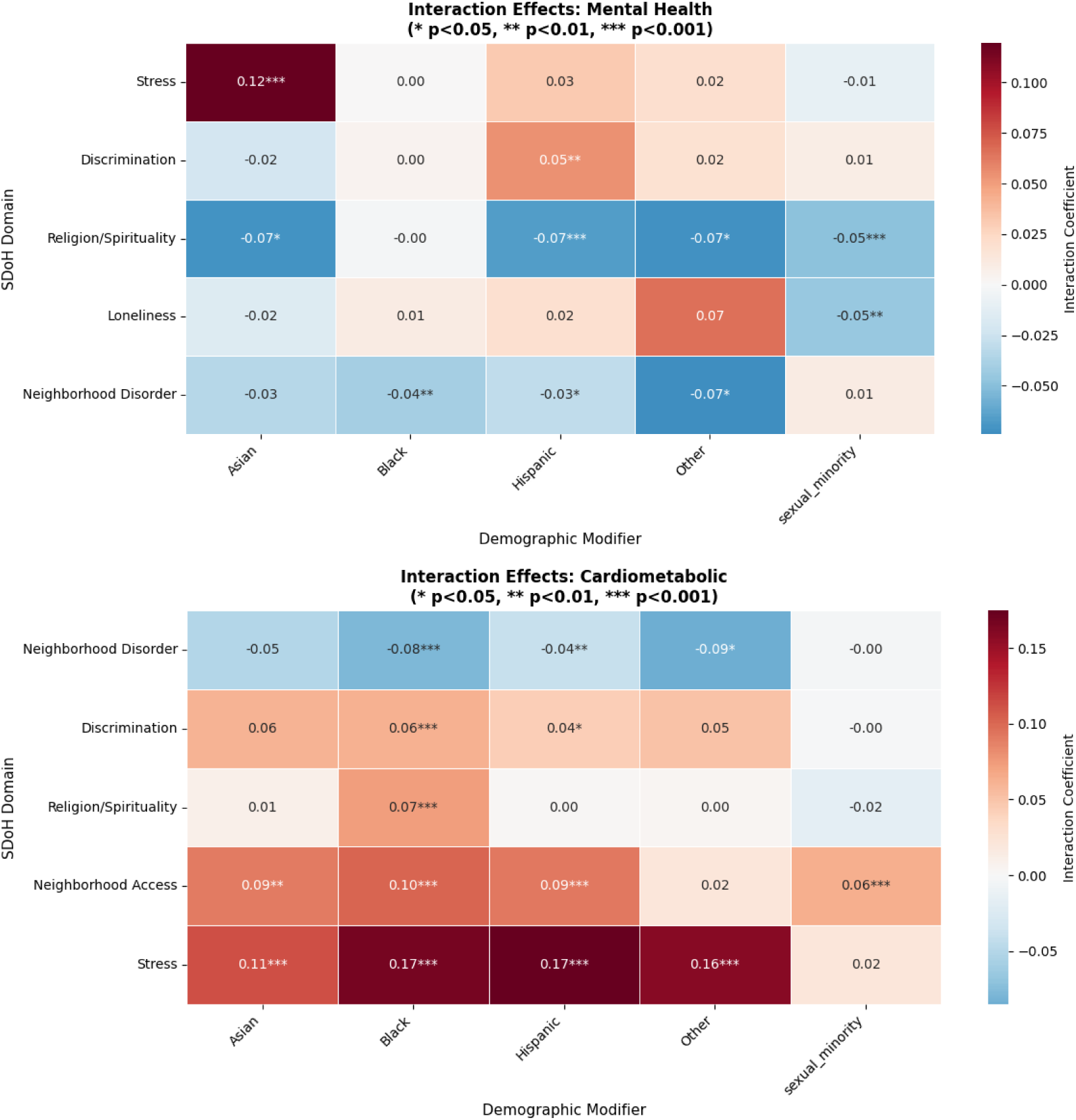
Interaction Coefficient Heatmaps. Heatmaps of interaction coefficients between SDoH domains (rows) and demographic modifiers (columns) for Mental Health (top) and Cardiometabolic (bottom) outcomes. Asterisks indicate significance levels (* p < 0.05, ** p < 0.01, *** p < 0.001).

**Fig 6: Fig F in S1 Text.**
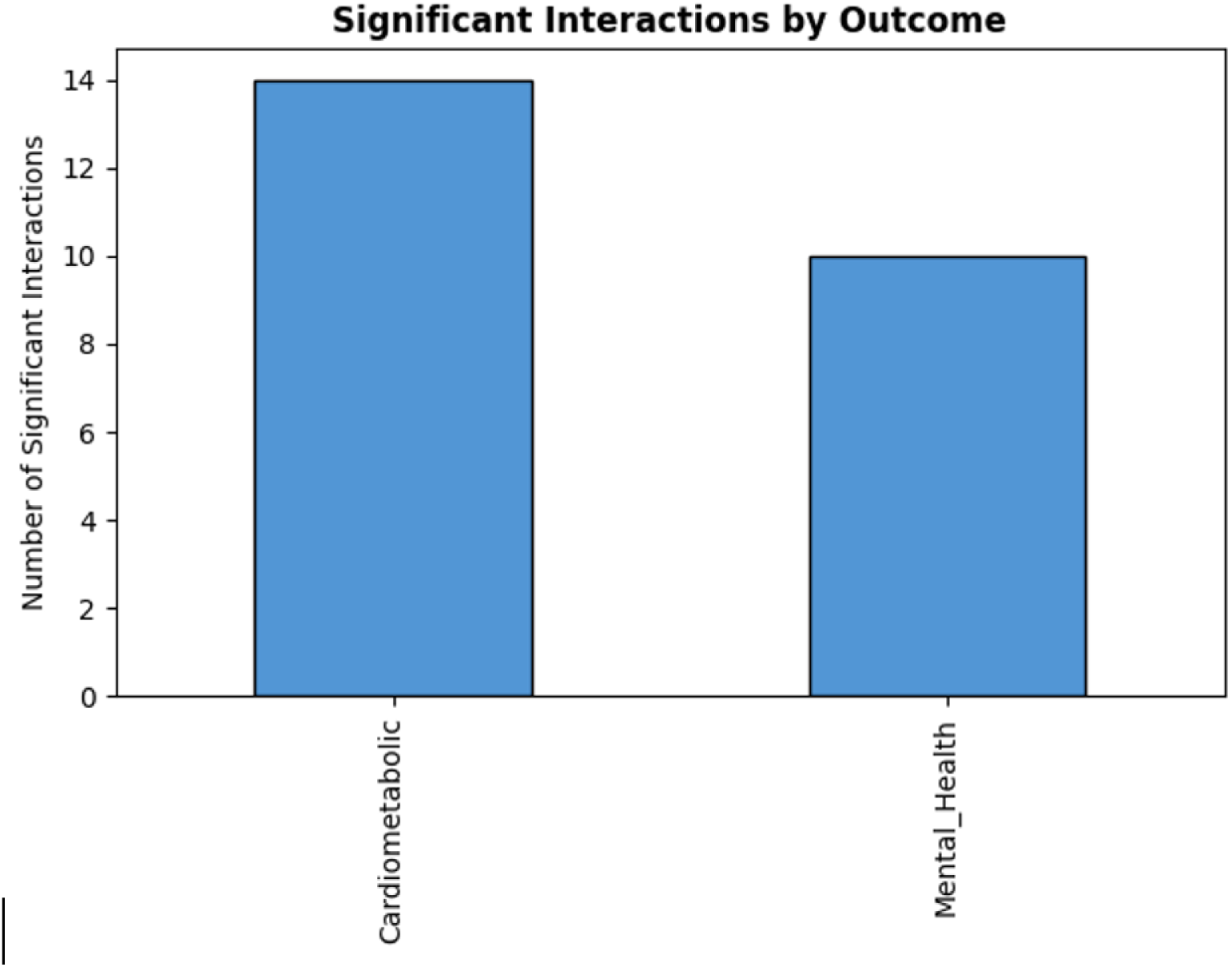
Significant Interactions by Outcome. Count of significant SDoH × demographic interactions by outcome cluster.

**Fig 7: Fig G in S1 Text.**
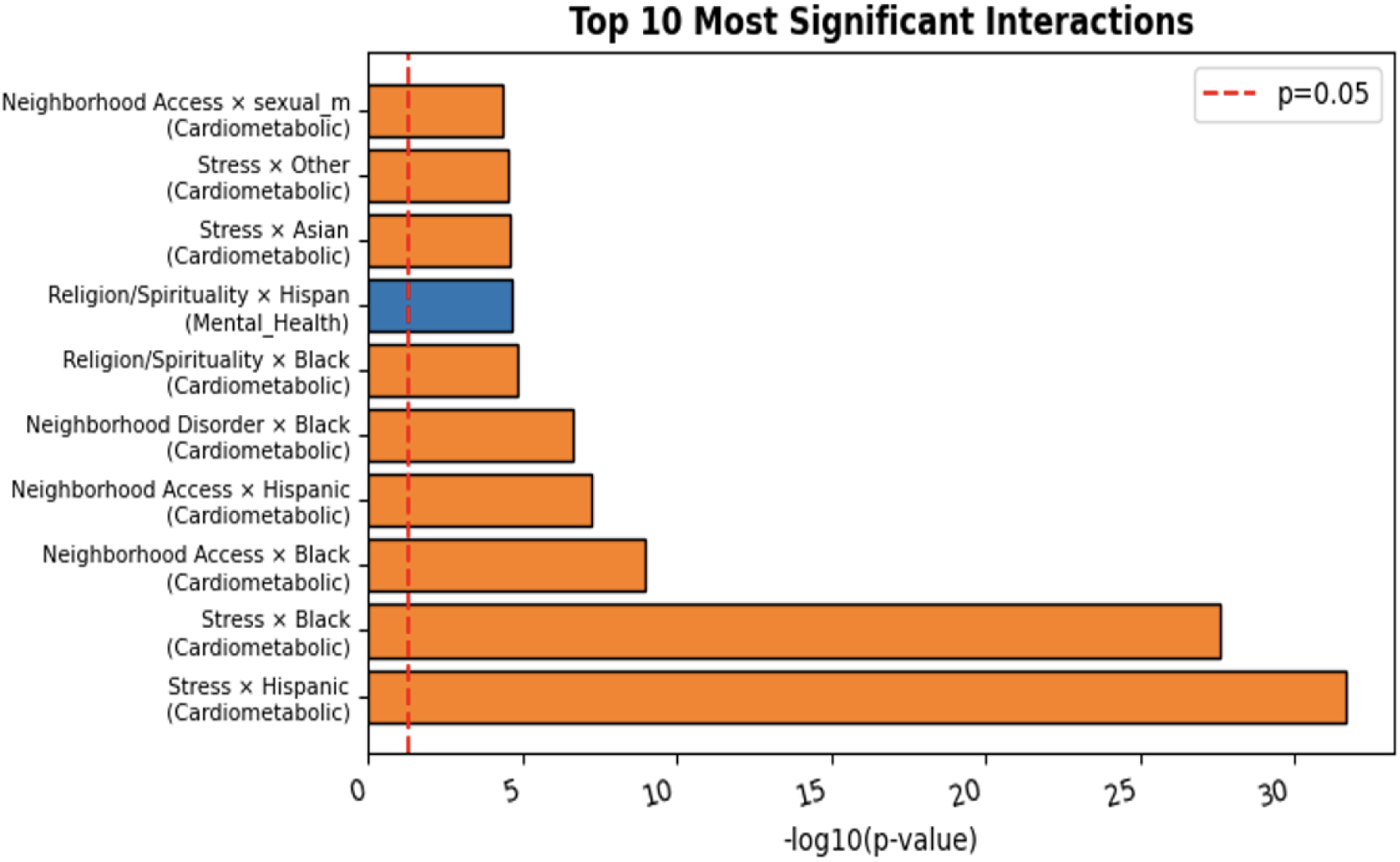
Top 10 Most Significant Interactions. SDoH × demographic interactions ranked by — log_10_(*p*-value). The dashed red line indicates the *p* = 0.05 threshold.

## Notes

### Competing Interest Statement

The authors have declared no competing interest.

### Funding Statement

The author(s) received no specific funding for this work.

### Author Declarations

This study was conducted using data from the All of Us Research Program. All participants provided informed consent for research use of their data as part of enrollment in the program. The All of Us Research Program is approved by the All of Us Institutional Review Board (IRB) at Vanderbilt University Medical Center. This specific analysis was conducted under the All of Us Registered Tier Data Access policies, which permit secondary analysis of de-identified data for approved research purposes.

